# Comparative Immunotherapeutic Strategies in Advanced Melanoma: A Systematic Review and Bayesian Meta-analysis of TIL and Engineered Viral Vector Therapies

**DOI:** 10.64898/2026.05.26.26353583

**Authors:** Joshua Anyachor

**Affiliations:** Faculty of Medicine and Health Sciences, University of Buckingham, Buckingham, United Kingdom

**Author notes:** Corresponding author: Joshua Anyachor Faculty of Medicine and Health Sciences, University of Buckingham, Buckingham, United Kingdom ORCID: 0009-0007-7137-6418.

**Keywords:** Melanoma, tumor-infiltrating lymphocytes, oncolytic viruses, engineered viral vectors, immunotherapy, Bayesian meta-analysis

## Abstract

Melanoma remains a treatment-refractory malignancy in a subset of patients despite major advances with immune checkpoint inhibitors. Tumor-infiltrating lymphocyte (TIL) therapy and engineered viral vector immunotherapies represent mechanistically distinct strategies for advanced melanoma, but their comparative clinical roles remain incompletely defined.

This systematic review and Bayesian meta-analysis evaluated clinical outcomes of TIL therapy and engineered viral vector immunotherapies in adults with unresectable stage III or IV melanoma. Searches of PubMed, Embase, Scopus, Web of Science, ClinicalTrials.gov, and grey literature sources identified 12 contemporary eligible studies published between 2015 and 2025. One additional pre-2015 pilot study was retained for qualitative historical context because it represented early clinical feasibility of viral vector-based immunotherapy in melanoma. Overall, 13 studies were included in the qualitative review, including four randomized controlled trials and nine single-arm studies, while eight studies were eligible for Bayesian quantitative synthesis of ORR.

TIL therapy demonstrated substantial standalone activity, particularly in checkpoint-exposed or PD-1-refractory populations, whereas viral vector therapies showed variable monotherapy activity and stronger responses when combined with immune checkpoint inhibition. The pooled Bayesian ORR estimate was 37.8% (95% highest density interval [HDI]: 30.6%-45.3%). Sensitivity analysis excluding the smallest included study yielded a similar pooled estimate of 38.3% (95% HDI: 30.4%-46.2%). Certainty of evidence was moderate for ORR and low for survival and safety outcomes due to heterogeneity, sparse reporting, and inconsistent endpoint definitions.

These findings support complementary rather than competing roles for TIL and engineered viral vector immunotherapies and highlight the need for biomarker-guided sequencing studies in advanced melanoma.

## Introduction

Melanoma is an aggressive and highly immunogenic malignancy that, despite accounting for less than 1% of skin cancers, is responsible for the majority of skin cancer–related deaths [1]. While early-stage disease is often curable with surgical excision, advanced melanoma remains therapeutically challenging, with five-year survival rates remaining below 30% in metastatic settings [2]. Historically, systemic chemotherapy yielded limited survival benefit and considerable toxicity, driving the transition toward more durable and immune-directed therapeutic strategies.

This transition led to the widespread adoption of immune checkpoint inhibitors (ICIs), particularly anti–PD-1 and anti–CTLA-4 therapies, which restore antitumor T-cell activity by reversing inhibitory immune signaling. Although ICIs have transformed the management of advanced melanoma and can produce durable long-term remissions, primary resistance and acquired relapse remain major clinical challenges, with only approximately 40–50% of patients achieving sustained responses [1–3]. Mechanisms underlying therapeutic resistance include impaired antigen presentation, T-cell exclusion, interferon signaling dysregulation, and upregulation of immunosuppressive ligands such as PD-L1, collectively contributing to an immune-evasive tumor microenvironment that limits the durability of checkpoint blockade.

In response to these limitations, two emerging immunotherapeutic strategies have gained increasing clinical and translational interest: tumor-infiltrating lymphocyte (TIL) therapy and engineered viral vector–based immunotherapies. TIL therapy involves the isolation and ex vivo expansion of autologous tumor-reactive lymphocytes followed by reinfusion after lymphodepleting conditioning. Because these lymphocytes are preselected within the tumor microenvironment, TIL therapy has demonstrated substantial efficacy in heavily pretreated and checkpoint-refractory melanoma populations [4]. However, broader implementation remains constrained by manufacturing complexity, treatment-associated toxicity, and logistical barriers related to individualized cell production.

Engineered viral vector immunotherapies offer a mechanistically distinct approach centered on in situ immune activation. Agents such as talimogene laherparepvec (T-VEC) selectively replicate within tumor tissue while expressing immunostimulatory cytokines including granulocyte–macrophage colony-stimulating factor (GM-CSF), thereby promoting local oncolysis and systemic antitumor immune priming [5]. In combination with ICIs, viral vectors may enhance antigen presentation, increase intratumoral immune infiltration, and convert immunologically “cold” tumors into more inflamed and treatment-responsive microenvironments [6].

Despite their mechanistic differences, TIL therapy and engineered viral vector immunotherapies converge on a shared therapeutic objective: overcoming immune resistance and restoring durable antitumor immunity in advanced melanoma. Importantly, these approaches may occupy complementary rather than competing roles within evolving melanoma treatment paradigms, particularly in PD-1–refractory disease where optimal sequencing and integration strategies remain poorly defined. Nevertheless, existing reviews have largely evaluated these modalities independently, and comparative quantitative syntheses directly evaluating their clinical outcomes across both randomized and prospective single-arm studies remain limited.

Accordingly, this study evaluates the comparative clinical outcomes of TIL therapy and engineered viral vector immunotherapies in advanced melanoma through a systematic review supplemented by Bayesian meta-analysis. By integrating evidence across heterogeneous trial designs, this analysis aims to estimate pooled therapeutic response, characterize between-study variability, and inform the evolving role of cellular and viral immunotherapies within biomarker-guided and combination-based melanoma treatment strategies.

## Methods

### Protocol and registration

This systematic review was conducted in accordance with the Cochrane Handbook for Systematic Reviews of Interventions [7] and reported following PRISMA 2020 guidelines [8] where applicable. Although the protocol was not prospectively registered in PROSPERO or a similar repository, all methodological decisions including eligibility criteria, search strategy, data extraction, risk of bias assessment, and synthesis approach were pre-specified and applied consistently. The absence of formal registration is acknowledged as a limitation and reflects the evolving scope of the research question during the initial scoping phase.

### Study eligibility and inclusion criteria

Eligibility criteria were defined using the PICO framework and focused on adults (≥18 years) with unresectable stage III or IV melanoma. Eligible interventions included autologous tumor-infiltrating lymphocyte (TIL) therapy and engineered viral vector based immunotherapies, administered as monotherapy or in combination with standard treatments. Comparator arms were not mandatory; when present, they included checkpoint inhibitors, chemotherapy, placebo, or no treatment in single-arm trials.

Studies evaluating genetically engineered T-cell receptor (TCR) therapies were excluded to maintain intervention comparability.

Eligible study designs included randomized controlled trials, phase I–III clinical trials, and clearly defined prospective single-arm studies. Preclinical studies, reviews, case reports, and editorials were excluded. Primary outcomes were objective response rate (ORR), progression-free survival (PFS), overall survival (OS), and grade ≥3 treatment-related adverse events.

Screening and extraction were conducted by a single reviewer using predefined eligibility criteria and standardized extraction forms. Although this deviates from PRISMA and Cochrane recommendations for dual independent review, structured eligibility criteria and standardized extraction forms were used to mitigate bias. This limitation is acknowledged and considered in the interpretation of findings.

**Table 1.** Study eligibility criteria based on the PICO framework.

| Component | Description |
| --- | --- |
| Population | Adults ($\geq 18$ years) with unresectable stage III or IV melanoma |
| Intervention | Autologous tumor-infiltrating lymphocyte (TIL) therapy or engineered viral vector therapy |
| Comparator | Any comparator (e.g., standard care, checkpoint inhibitors), or none for single-arm trials |
| Outcomes | Objective response rate (ORR), progression-free survival (PFS), overall survival (OS), treatment-related adverse events |

### Information Sources and Search Strategy

A comprehensive literature search was performed on March 16, 2025, across PubMed (MEDLINE), Embase, Scopus, and Web of Science. Grey literature sources, including the Melanoma Research Alliance and ClinicalTrials.gov, were also reviewed to capture unpublished or ongoing trials and minimize publication bias.

The search covered studies published from January 1, 2015, and March 31, 2025. Medical Subject Headings (MeSH) + free-text terms were combined using Boolean operators. The full database-specific search strategies, including Boolean operators, field tags, and date limits, are provided in Appendix A7. Key terms included melanoma, tumor-infiltrating lymphocytes, oncolytic viruses, gene therapy, TIL therapy, engineered vectors, and immune modulation. Duplicates were removed using Zotero reference management software, and remaining records were screened against predefined eligibility criteria. The study selection process is summarized in the PRISMA flow diagram (Figure 1).

**Fig. 1.**
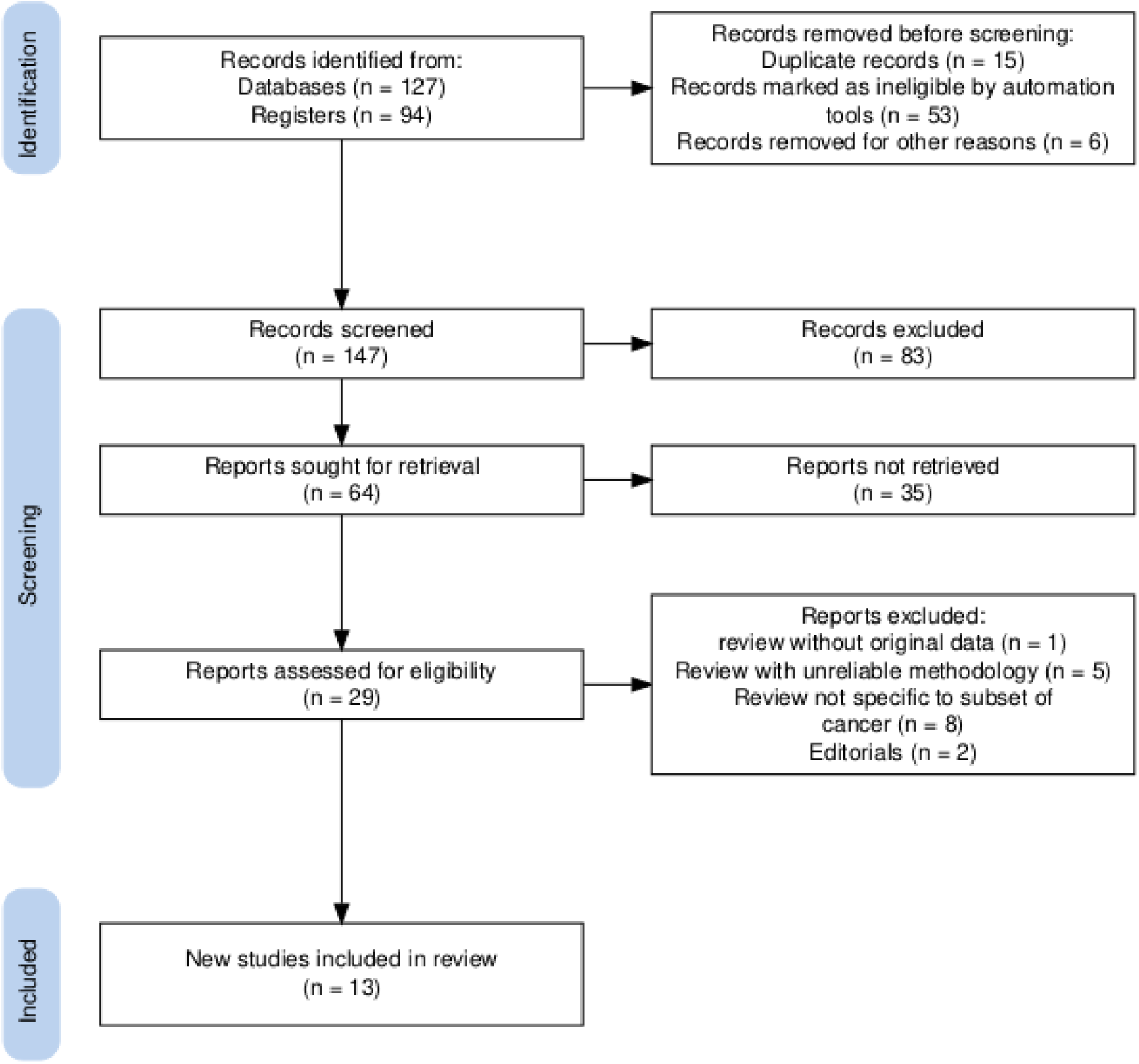
PRISMA flow diagram of study selection. Flow of records through the study selection process, detailing numbers identified, screened, excluded, and included. Diagram generated using the PRISMA 2020 flow diagram tool.

**Fig. 2.**
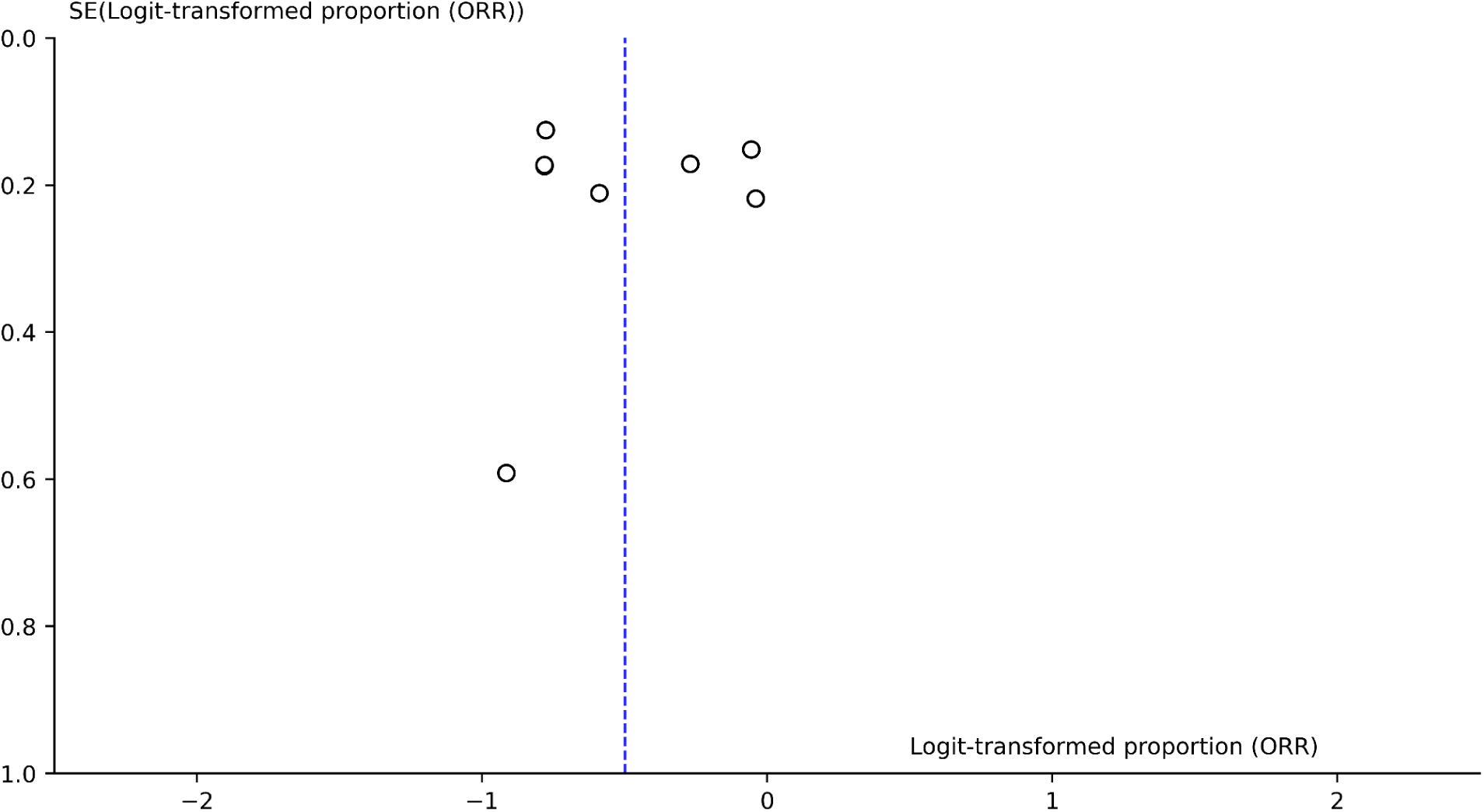
Funnel plot of logit-transformed objective response rates (ORRs) versus standard error. Each point represents one study included in the Bayesian quantitative synthesis. Distribution was visually assessed for asymmetry; no funnel plot boundaries were applied. Interpretation is limited by the small number of studies and substantial heterogeneity. Generated using PyMC3 and Matplotlib.

One pre-2015 pilot study, Mastrangelo et al. (1999), was retained as a historical contextual study because it represented early clinical feasibility of vaccinia/GM-CSF viral vector immunotherapy in melanoma. This study was included only in the qualitative synthesis and study-characteristics tables to contextualize the development of engineered viral vector approaches. It was excluded from the Bayesian quantitative synthesis and did not contribute to pooled ORR estimates, sensitivity analyses, meta-regression, or GRADE certainty judgments for contemporary efficacy evidence.

### Data extraction

A standardized, piloted extraction form was used to ensure consistent data collection. Extracted variables included study design, sample size, trial phase, clinical setting, geographic location, and funding source. Intervention-level details included TIL expansion protocols, viral vector type, use of IL-2, and combination regimens. Patient-level data included demographics, disease stage, and prior exposure to PD-1 or CTLA-4 inhibitors.

Clinical outcomes included ORR, PFS, OS, duration of response, and grade ≥3 treatment-related adverse events. Where reported, immunologic correlates (e.g., cytokine levels, T-cell infiltration, immune checkpoint expression) were also recorded. Supplementary data were obtained from appendices, registries, or related publications when necessary. No imputation of missing values was performed.

### Risk of bias assessment

Risk of bias was assessed using the Cochrane RoB 2 tool for randomized controlled trials and ROBINS-I for non-randomized or single-arm studies. RoB 2 evaluated bias across five domains: randomization, deviations from intended interventions, missing outcome data, outcome measurement, and selection of reported results. ROBINS-I assessed confounding, participant selection, intervention classification, missing data, and outcome measurement.

All assessments were conducted using standardized criteria, and justifications were documented where ratings were uncertain. Bias ratings were not incorporated directly into the Bayesian meta-analysis but informed interpretation and GRADE certainty assessments. Summary judgments are presented in the Results section, with full domain-level ratings provided in Appendices A3 and A4.

### Effect measures

The primary outcome was objective response rate (ORR), defined as the proportion of patients achieving complete or partial tumor response. ORR was prioritized due to its consistent reporting across studies, applicability to both randomized and single-arm designs, and reduced susceptibility to censoring and variable follow-up durations.

To stabilize variance and minimize the influence of extreme proportions, individual study ORRs were logit-transformed prior to meta-analysis. A Bayesian random effects model was used to estimate pooled ORR, with results reported as 95 % highest density intervals (HDIs). Due to inconsistent definitions and insufficient reporting, PFS and OS were not pooled and were summarized narratively.

### Data synthesis and statistical analysis

Both qualitative and quantitative syntheses were conducted. Study characteristics and clinical outcomes were summarized descriptively and in tabular form.

A Bayesian random-effects meta-analysis was used to estimate pooled ORR among studies meeting criteria for quantitative synthesis. Although 13 studies were included in the overall systematic review, only 8 studies with sufficiently comparable interventions and complete ORR reporting were included in the Bayesian quantitative analysis. A Bayesian framework was selected to accommodate heterogeneous study designs, account for uncertainty in sparse and small-sample evidence settings, and provide probabilistic interpretation of treatment effects.

Exploratory meta-regression using the metafor package in R evaluated whether study design (RCT vs single-arm), therapeutic modality (TIL vs viral vector), or treatment context (monotherapy vs combination) moderated ORR. Sensitivity analyses included leave-one-out testing, exclusion of small sample studies (N < 20), restriction to randomized trials, and removal of statistical outliers. Full analytical code used for the Bayesian meta-analysis and exploratory meta-regression is provided in Appendices A5 and A6.

The primary Bayesian model included eight studies, and a sensitivity analysis excluding the small-sample Cui et al. study [10] was performed to evaluate the influence of small-study inclusion on the pooled ORR estimate.

Weakly informative priors were used: a normal(0, 10) prior for the overall logit effect size (μ) and a half-normal(0, 1) prior for between-study standard deviation (τ), consistent with recommendations for sparse data.

Between-study heterogeneity was quantified using the between-study standard deviation (τ). Exploratory frequentist pooling using RevMan yielded an I² of 77 %, indicating moderate-to-high heterogeneity. Convergence diagnostics supported model reliability, with R-hat values < 1.01, no divergent transitions, and satisfactory chain mixing.

Small-study effects were explored using scatterplots of logit-transformed ORR against study standard error (SE) and Egger’s regression test. Funnel plot boundaries and trim-and-fill analyses were not performed due to the limited number of studies and substantial heterogeneity.

Not all included studies (n = 13) were eligible for quantitative synthesis. Studies were excluded from the Bayesian meta-analysis if they did not meet the predefined comparability criteria for pooled ORR estimation or lacked sufficient data to compute logit-transformed ORRs. Eight studies with sufficiently comparable interventions and complete ORR reporting were included in the primary Bayesian synthesis, including Cui et al. [10]. A sensitivity analysis excluding this small-sample study (N = 14) was additionally performed to evaluate its influence on the pooled ORR estimate. The qualitative synthesis therefore included all eligible studies, whereas the quantitative analysis was restricted to studies meeting predefined criteria for comparability and outcome reporting.

### Certainty assessment

Certainty of evidence was assessed using the GRADE framework [9], evaluating risk of bias, inconsistency, indirectness, imprecision, and publication bias. Publication bias was assessed using scatterplots and Egger’s test (p = 0.081), with no significant asymmetry observed.

ORR was rated moderate certainty, while PFS, OS, and safety outcomes were rated low certainty due to heterogeneity, inconsistent definitions, and incomplete reporting.

### Ethical considerations

All included studies were peer-reviewed clinical trials conducted with ethical approval and informed consent. This review involved secondary analysis of publicly available data and therefore did not require additional ethical approval.

## Results

### Study selection

A total of 221 records were identified through database and trial registry searches. Before screening, 74 records were removed, including 15 duplicate records, 53 records marked as ineligible by automation tools, and 6 records removed for other reasons. This left 147 records for title and abstract screening, of which 83 were excluded. Sixty-four reports were sought for retrieval, and 35 were not retrieved. Twenty-nine full-text reports were assessed for eligibility, of which 16 were excluded: 1 review without original data, 5 reviews with unreliable methodology, 8 reports not specific to the relevant cancer subset, and 2 editorials. Thirteen studies met the inclusion criteria and were included in the systematic review.

### Study characteristics

The included studies comprised four randomized controlled trials and nine single-arm trials evaluating tumor-infiltrating lymphocyte (TIL) therapy and engineered viral vector-based immunotherapies in advanced melanoma.

Geographic distribution and funding sources are summarized in Tables 2 and 3.

**Table 2.** Geographic Characteristics of Included Studies.

| Study | Country / Region | Multicenter? | Trial Type | Notes |
| --- | --- | --- | --- | --- |
| Rohaani et al. (2022) | Netherlands & Denmark | Yes | Phase 3 RCT | Point-of-care TIL therapy |
| Chesney et al. (2022)<br>[T-VEC + pembro] | Global (USA, EU, Asia) | Yes | Phase 3 RCT | Amgen-sponsored combination trial |
| Chesney et al. (2023)<br>[T-VEC + ipi] | USA & EU | Yes | Phase 2 RCT | T-VEC + ipilimumab |
| Andtbacka et al. (2019) | USA, UK, EU | Yes | Phase 3 RCT | OPTiM trial |
| Chesney et al. (2022,<br>Lifileucel) | USA & Europe | Yes | Phase 2, single-arm | Iovance-sponsored centralized TIL trial |
| Nguyen et al. (2019) | Canada | No | Phase 2, single-arm | Real-world IL-2 + TIL study |
| van den Berg et al. (2020) | Netherlands | No | Phase 1/2, single-arm | Pilot TIL trial at NKI |
| Seitter et al. (2021) | USA | No | Observational,<br>single-arm | Real-world PD-1 stratified cohort |
| Cui et al. (2022) | China (Beijing) | No | Phase 1b, single-arm | OrienX010 oncolytic vector |
| Wong et al. (2024) | USA | No | Phase 2, single-arm | RP1 + nivolumab |
| Sussman et al. (2024) | USA (Boston) | No | Phase 2, single-arm | Autologous GM-CSF vaccine |
| Yamazaki et al. (2022) | Japan | Yes | Phase 1/2, single-arm | T-VEC trial in Japanese cohort |
| Mastrangelo et al. (1999) | USA | No | Pilot, single-arm | Early vaccinia-based vector trial |
| **Note:** Summarizes geographic origin, multicenter status, and trial type. Regions are based on reported recruitment sites. “Multicenter” indicates recruitment across multiple hospitals or institutions. Geographic distribution informs generalizability across populations and healthcare systems. |  |  |  |  |

**Table 3.** Funding and Sponsorship of Included Studies.

| Study | Funding Source | Industry Sponsored? | Notes |
| --- | --- | --- | --- |
| Rohaani et al. (2022) | Dutch Cancer Society / EU | No | Public academic funding |
| Chesney et al. (2022) [T-VEC + pembro] | Amgen | Yes | Amgen-developed vector |
| Chesney et al. (2023) [T-VEC + ipi] | Amgen | Yes | Sponsor-funded combination |
| Andtbacka et al. (2019) | Amgen | Yes | OPTiM trial sponsor |
| Chesney et al. (2022, Lifileucel) | Iovance Biotherapeutics | Yes | Lifileucel product trial |
| Nguyen et al. (2019) | Not stated | Likely No | Academic hospital-based |
| van den Berg et al. (2020) | Netherlands Cancer Institute | No | Non-commercial pilot |
| Seitter et al. (2021) | NIH / Academic | No | Public real-world study |
| Cui et al. (2022) | Seven and Eight Biopharma | Yes | Industry-developed HSV-1 |
| Wong et al. (2024) | Replimune | Yes | RP1 proprietary vector |
| Sussman et al. (2024) | Dana-Farber / NIH | No | Academic vaccine study |
| Yamazaki et al. (2022) | Amgen | Yes | Japan-based Amgen trial |
| Mastrangelo et al. (1999) | Not stated | Possibly | Unclear historical funding |
| **Note:** Outlines declared funding sources and sponsorship status. “Industry sponsored” refers to support from pharmaceutical or biotech companies. Public or academic sponsorship includes government, nonprofit, or institutional funding. Unreported funding is based on available disclosures. |  |  |  |

### Risk of Bias Assessment

Risk of bias was assessed for each included study. Most randomized controlled trials were judged to have low risk of bias, whereas single-arm studies showed moderate to serious concerns, primarily due to confounding and lack of blinding.

**Table 4.** Risk of bias assessment for randomized controlled trials.

| Study | Randomization process | Deviations of intended interventions | Missing outcome data | Outcome measurement | Selection of the reported result | Overall risk |
| --- | --- | --- | --- | --- | --- | --- |
| Andtbacka et al., 2019 | Low risk | Low risk | Low risk | Low risk | Low risk | Low |
| Rohaani et al. (2022) | Low risk | Low risk | Low risk | Low risk | Low risk | Low |
| Chesney et al. (2022) | Low risk | Low risk | Some concerns | Low risk | Some concerns | Some concerns |
| Chesney et al. (2023) | Low risk | Low risk | Low risk | Low risk | Low risk | Low |
| *Note : Risk of bias was assessed using the Cochrane Risk of Bias 2 (RoB 2) tool. “Low” indicates low risk of bias across all domains; “some concerns” indicates potential bias in at least one domain; and “high” indicates high risk of bias that may substantially lower confidence in the results.* |  |  |  |  |  |  |

**Table 5.** Risk of bias assessment for non-randomized and single-arm studies (ROBINS-I).

| Study | Confounding | Participant selection | Intervention Classification | Deviations from intended interventions | Missing data | Outcome measurement | Selection of reported result | Overall risk |
| --- | --- | --- | --- | --- | --- | --- | --- | --- |
| Chesney et al. (2022) [Lifileucel] | Moderate risk | Low risk | Low risk | Low risk | Moderate risk | Low risk | Moderate risk | Moderate risk |
| Nguyen et al. (2019) | Serious risk | Moderate risk | Low risk | Low risk | Serious risk | Moderate risk | moderate risk | Serious risk |
| van den Berg et al. (2020) | Serious risk | Moderate risk | Low risk | Low risk | Moderate risk | Moderate risk | Moderate risk | Serious risk |
| Seitter et al. (2021) | Moderate risk | Low risk | Low risk | Low risk | Moderate risk | Low risk | Low risk | Moderate risk |
| Cui et al. (2022) | Serious risk | moderate risk | Low risk | Low risk | Moderate risk | Moderate risk | Moderate risk | serious risk |
| Wong et al. (2024) | Moderate risk | Low risk | Low risk | Low risk | Moderate risk | Moderate risk | Moderate risk | Moderate risk |
| Mastrangelo et al. (1999) | Serious risk | Serious risk | Low risk | Low risk | Moderate risk | Low risk | Moderate risk | Serious risk |
| Sussman et al. (2024) | serious risk | Moderate risk | Low risk | Low risk | Moderate risk | Moderate risk | Serious risk | Serious risk |
| Yamazaki et al. (2022) | Moderate risk | Low risk | Low risk | Low risk | Low risk | Low risk | Low risk | Moderate risk |
| *Note: Risk of bias was assessed using the ROBINS-I tool. “Low,” “moderate,” and “serious” indicate increasing risk of bias that may reduce confidence in the findings.* |  |  |  |  |  |  |  |  |

### Outcomes

Studies reported objective response rate (ORR), progression-free survival (PFS), overall survival (OS), and grade ≥3 adverse events. Outcomes are summarized by therapy type (TIL vs. vector-based) and study design (randomized vs. single-arm).

### TIL therapies randomized controlled trials

**Table 6.**
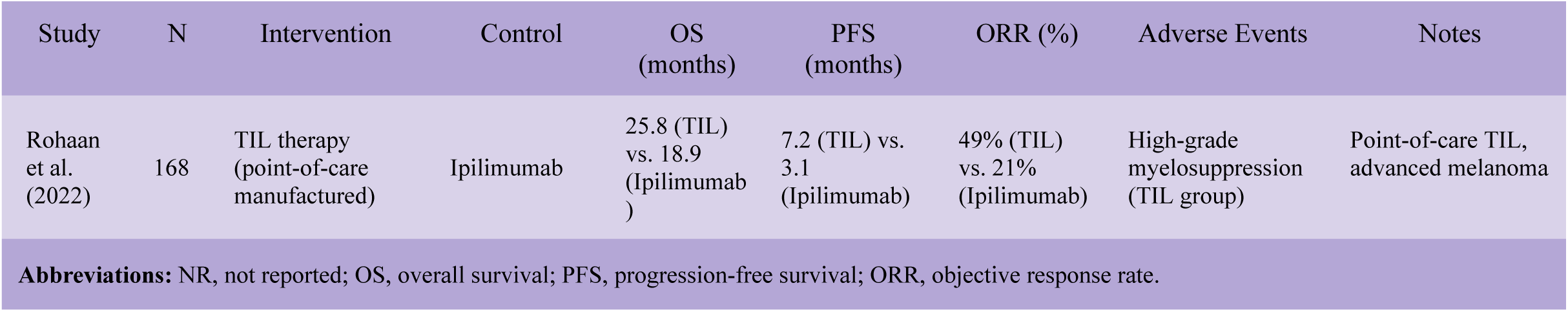
Randomized controlled trials of TIL therapy.

| Study | N | Intervention | Control | OS (months) | PFS (months) | ORR (%) | Adverse Events | Notes |
| --- | --- | --- | --- | --- | --- | --- | --- | --- |
| Rohaani et al. (2022) | 168 | TIL therapy (point-of-care manufactured) | Ipilimumab | 25.8 (TIL) vs. 18.9 (Ipilimumab) | 7.2 (TIL) vs. 3.1 (Ipilimumab) | 49% (TIL) vs. 21% (Ipilimumab) | High-grade myelosuppression (TIL group) | Point-of-care TIL, advanced melanoma |
| **Abbreviations:** NR, not reported; OS, overall survival; PFS, progression-free survival; ORR, objective response rate. |  |  |  |  |  |  |  |  |

### TIL therapies single-arm studies

**Table 7.** Single-arm clinical trials of TIL therapy.

| Study | N | Intervention | OS (months) | PFS (months) | ORR (%) | Adverse Events | Notes |
| --- | --- | --- | --- | --- | --- | --- | --- |
| Chesney et al. (2022) | 153 | Lifileucel (centralized TIL therapy) | 13.9 | 4.1 | 31.4% (8 CR, 40 PR) | Thrombocytopenia (76.9%), anemia (50.0%), febrile neutropenia (41.7%) | Heavily pretreated metastatic melanoma |
| (Nguyen et al., 2019) | 12 | TIL + Low-dose IL-2 | NR | NR | 16.7% (2 PRs) | Expected, manageable; 1 unconfirmed PR; 6 SD | Small exploratory study |
| van den Berg et al. (2020) | 10 | Autologous TIL Therapy | NR | NR | 50% (2 CR, 3 PR) | High-dose IL-2 toxicities; ICU care in 1 case | Personalized TIL, very small cohort |
| (Seitter et al., 2021). ongoing-trial | 139 | Autologous TIL + NMA + IL-2 | Median OS: 28.5 (PD-1 naïve); 11.6 (PD-1 refractory) | Not reported | 43.3% | IL-2 toxicity; myelosuppression; expected profile | Real-world TIL data, stratified |
| **Abbreviations:** NR, not reported; OS, overall survival; PFS, progression-free survival; ORR, objective response rate; PR, partial response; NMA, non-myeloablative conditioning. |  |  |  |  |  |  |  |

### Engineered viral vector therapies randomized controlled trials

**Table 8.**
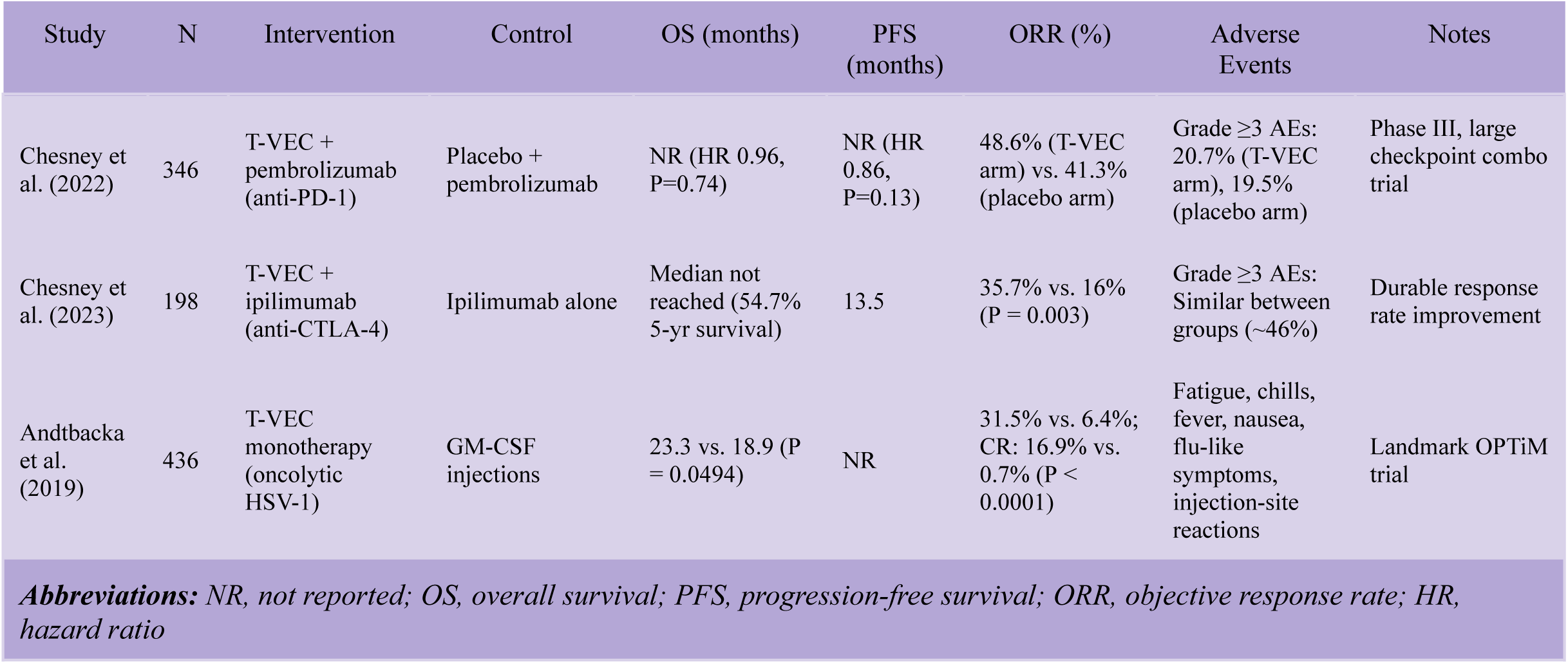
Randomized controlled trials of engineered viral vector therapies.

| Study | N | Intervention | Control | OS (months) | PFS (months) | ORR (%) | Adverse Events | Notes |
| --- | --- | --- | --- | --- | --- | --- | --- | --- |
| Chesney et al. (2022) | 346 | T-VEC + pembrolizumab (anti-PD-1) | Placebo + pembrolizumab | NR (HR 0.96, P=0.74) | NR (HR 0.86, P=0.13) | 48.6% (T-VEC arm) vs. 41.3% (placebo arm) | Grade $\geq 3$ AEs: 20.7% (T-VEC arm), 19.5% (placebo arm) | Phase III, large checkpoint combo trial |
| Chesney et al. (2023) | 198 | T-VEC + ipilimumab (anti-CTLA-4) | Ipilimumab alone | Median not reached (54.7% 5-yr survival) | 13.5 | 35.7% vs. 16% (P = 0.003) | Grade $\geq 3$ AEs: Similar between groups (~46%) | Durable response rate improvement |
| Andtbacka et al. (2019) | 436 | T-VEC monotherapy (oncolytic HSV-1) | GM-CSF injections | 23.3 vs. 18.9 (P = 0.0494) | NR | 31.5% vs. 6.4%; CR: 16.9% vs. 0.7% (P < 0.0001) | Fatigue, chills, fever, nausea, flu-like symptoms, injection-site reactions | Landmark OPTiM trial |

**Abbreviations:** NR, not reported; OS, overall survival; PFS, progression-free survival; ORR, objective response rate; HR, hazard ratio

**Table 9.** Single-Arm Trials.

| Study | N | Intervention | OS (months) | PFS (months) | ORR (%) | Adverse Events | Notes |
| --- | --- | --- | --- | --- | --- | --- | --- |
| Cui et al. (2022) | 14 | OrienX010 (oncolytic HSV-1) | 17.4 | 3.0 | 28.6 | Fever, injection site reaction; 1 grade $\geq 3$ | China-based phase I study |
| Wong et al (2024) | 156 | RP1 + Nivolumab | DoR>24 mo; 78% ongoing | Not reported | 31.4% | Mostly grade 1-2, 1 grade 5 (myocarditis from nivolumab | Checkpoint-refractory cohort |
| Mastrangelo et al., 1999 | 7 | Vaccinia/GM-CSF | NR | NR | 28.6% | Mostly mild flu-like symptoms (12%) inflammation; no toxicity | Early viral vector feasibility |
| Sussman et al., 2024 | 61 | Autologous GM-CSF Vaccine | Stage III: 50.7; stage IV: 21.3 | Stage III: 11.5; stage IV: 3.0 | 1.6% | mild local erythema(57%),fatigue(26%),nausea(10%); 1 serious AE | Poor ORR but long OS in Stage III |
| Yamazaki et al., 2022 | 18 | T-VEC (oncolytic HSV-1) | 22.87 | 3.07 | 11.1% | Grade $\geq 3$ AEs: 27.8%; common: pyrexia, malaise | Japan-based cohort |

**Abbreviations:** NR, not reported; OS, overall survival; PFS, progression-free survival; ORR, objective response rate; AE, adverse event; DoR, duration of response.

### Overview of clinical outcomes

This systematic review included 13 clinical trials: four randomized controlled trials (RCTs) and nine single-arm studies. Five evaluated tumor-infiltrating lymphocyte (TIL) therapy and eight assessed engineered viral vector-based immunotherapies in advanced melanoma. The primary outcome across studies was objective response rate (ORR), with additional data reported for progression-free survival (PFS), overall survival (OS), and treatment-related adverse events.

### TIL-Based Therapies

Reported ORRs for TIL therapies ranged from 16.7% to 50% across included studies [11, 15–18]. The largest randomized controlled trial, Rohaan et al. [11], included 168 patients and reported an ORR of 49% for TIL therapy compared with 21% for ipilimumab, with median PFS of 7.2 months versus 3.1 months, respectively. In the pooled C-144-01 lifileucel analysis, Chesney et al. [15] reported an ORR of 31.4% and a median OS of 13.9 months among 153 patients with advanced melanoma previously treated with immune checkpoint inhibitors and targeted therapies. Seitter et al. [18] reported an ORR of 43.3% in 139 patients treated with autologous TIL therapy, with OS stratified by prior PD-1 exposure: 28.5 months in PD-1-naive patients versus 11.6 months in PD-1-refractory patients.

### Engineered Viral Vector Therapies

For vector-based therapies, reported ORRs ranged from 1.6% to 48.6%, with substantial variability across trial design, vector platform, and combination treatment context [10, 12–14, 19–22]. In the OPTiM trial, Andtbacka et al. [14] reported that T-VEC monotherapy achieved an ORR of 31.5% compared with 6.4% for GM-CSF. In randomized combination trials, T-VEC plus ipilimumab yielded an ORR of 35.7% compared with 16% for ipilimumab alone [13], while T-VEC plus pembrolizumab yielded an ORR of 48.6% compared with 41.3% for placebo plus pembrolizumab [12]. Smaller studies using HSV-based or related viral vector platforms reported ORRs of 28.6% for OrienX010 [10] and 31.4% for RP1 plus nivolumab [19]. One study evaluating a GM-CSF-secreting vaccine reported a notably low ORR of 1.6%, although longer OS was observed among patients with stage III disease [20].

### Subgroup and Sensitivity Analyses

Exploratory subgroup and sensitivity analyses were conducted to assess variability and test the robustness of pooled ORR estimates. TIL therapies tended to show higher response rates in PD-1–refractory populations, while vector-based therapies were most effective when combined with checkpoint inhibitors. Excluding small-sample studies or statistical outliers did not materially affect the pooled estimate. These analyses were exploratory and are summarized narratively.

### Bayesian Meta-Analysis of ORR and Certainty of Evidence (GRADE)

Of the 13 included studies, 8 studies with sufficiently comparable interventions and complete ORR reporting were included in the Bayesian quantitative synthesis, while all studies were retained in the qualitative synthesis. Objective response rate (ORR) was selected as the primary outcome for quantitative synthesis due to its consistent reporting across studies. A GRADE assessment rated the certainty of evidence for ORR as moderate due to heterogeneity and imprecision. Overall survival (OS), progression-free survival (PFS), and adverse events were rated low certainty due to inconsistent definitions, sparse data, and reporting variability. For visualization purposes, the forest plot displays studies included in the Bayesian synthesis together with logit-transformed ORR estimates and corresponding study-level confidence intervals.

**Table 10.** Certainty of evidence across outcomes.

| Studies Reporting Outcome | Number of Studies | Certainty of Evidence | Reasons for Downgrade |
| --- | --- | --- | --- |
| ORR | 12 contemporary studies reported ORR; 1 historical contextual study reported ORR qualitatively; 8 studies included in Bayesian synthesis | Moderate | Moderate heterogeneity, imprecise estimates (wide HDIs ) |
| OS | 10 | Low | High heterogeneity, inconsistent survival reporting |
| PFS | 8 | Low | Sparse data, substantial between-study variability |
| Adverse Events | 12 | Low | Inconsistent reporting, subjective classification of toxicity severity |
| **Note:** Certainty was assessed using the GRADE framework, which evaluates risk of bias, inconsistency, indirectness, imprecision, and publication bias. |  |  |  |

The Bayesian random-effects meta-analysis estimated a pooled ORR of 37.8% among the eight studies included in the quantitative synthesis (95% highest density interval [HDI]: 30.6%–45.3%). A sensitivity analysis excluding the small-sample Cui et al. study [10] yielded a similar pooled estimate of 38.3% (95% HDI: 30.4%–46.2%), supporting the robustness of the findings. Exploratory meta-regression identified no statistically significant effect modification by therapy type or study design. Between-study heterogeneity was moderate (τ ≈ 0.46), and Egger’s test showed no statistically significant evidence of small-study effects or publication bias (p = 0.081).

Forest plot estimates are shown in Figure 3, and the logit-transformed values used for Bayesian meta-analysis are presented in Table 11.

**Fig. 3.**
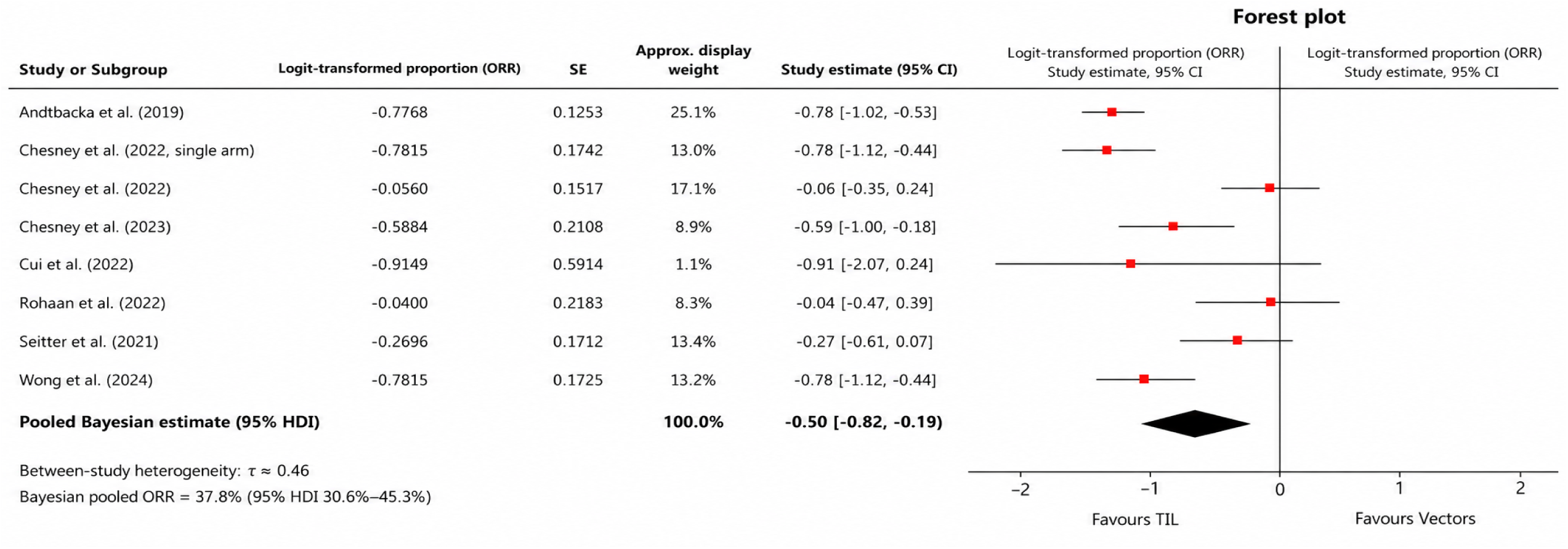
Forest plot of logit-transformed objective response rates (ORRs) for studies included in the Bayesian quantitative synthesis. Individual study estimates are represented by red square markers with horizontal study-level 95% confidence intervals. The pooled Bayesian estimate is shown as a black diamond with its corresponding 95% highest density interval (HDI). Between-study heterogeneity is summarized by τ. Generated using PyMC3 and Matplotlib.

**Table 11.** Input Data for Bayesian Meta-Analysis.

| Study | Design | Therapy | ORR(%) | N | Logit_ORR | SE_Logit_ORR | Included in Bayesian Synthesis |
| --- | --- | --- | --- | --- | --- | --- | --- |
| Rohaani et al. (2022) | RCT | TIL | 49 | 84 | -0.04 | 0.2183 | Yes |
| Chesney et al. (2022) | RCT | Vector (T-VEC + pembro) | 48.6 | 174 | -0.056 | 0.1517 | Yes |
| Chesney et al. (2023) | RCT | Vector (T-VEC + ipi) | 35.7 | 98 | -0.5884 | 0.2108 | Yes |
| Andtbacka et al. (2019) | RCT | T-VEC | 31.5 | 295 | -0.7768 | 0.1253 | Yes |
| Chesney et al. (2022, single-arm) | single-arm | TIL | 31.4 | 153 | -0.7815 | 0.1742 | Yes |
| Nguyen et al. (2019) | single-arm | TIL | 16.7 | 12 | -1.607 | 0.7740 | No |
| van den Berg et al. (2020) | single-arm | TIL | 50.0 | 10 | 0.0 | 0.6325 | No |
| Seitter et al. (2021) | single-arm | TIL | 43.3 | 139 | -0.2700 | 0.1712 | Yes |
| Mastrangelo et al. (1999) | single-arm | Vector (Vaccinia/GM-CSF) | 28.6 | 7 | -0.9149 | 0.8364 | No |
| Cui et al. (2022) | single-arm | Vector (OrienX010) | 28.6 | 14 | -0.9149 | 0.5914 | Yes |
| Wong et al. (2024) | single-arm | Vector (RP1 + nivo) | 31.4 | 156 | -0.7815 | 0.1725 | Yes |
| Sussman et al. (2024) | single-arm | Vector (GM-CSF vaccine) | 1.6 | 61 | -4.119 | 1.020 | No |
| Yamazaki et al. (2022) | single-arm | Vector (T-VEC) | 11.1 | 18 | -2.081 | 0.7503 | No |
| *Note: ORRs were logit-transformed, and standard errors were calculated using binomial variance approximations. The final column indicates whether each study was included in the Bayesian pooled estimate.* |  |  |  |  |  |  |  |

## Discussion

This systematic review supplemented by Bayesian meta-analysis identified broadly comparable objective response rates (ORRs) between tumor-infiltrating lymphocyte (TIL) therapies and engineered viral vector immunotherapies in advanced melanoma. Across included TIL studies, reported ORRs ranged from approximately 16.7% to 50%, with agents such as lifileucel demonstrating clinically meaningful activity in heavily pretreated and PD-1–exposed populations. These findings are consistent with prior evidence syntheses evaluating adoptive cellular therapy in melanoma [23].

Engineered viral vector–based therapies, including talimogene laherparepvec (T-VEC) and herpes simplex virus (HSV)-derived platforms, similarly demonstrated variable but clinically relevant antitumor activity, particularly when combined with immune checkpoint inhibitors [6, 12, 13] . Although posterior uncertainty remained substantial, pooled estimates were compatible with overlapping therapeutic activity across both modalities, and exploratory meta-regression did not identify significant effect modification by therapy type. Collectively, these findings support the interpretation that both TIL and viral vector immunotherapies may provide clinically meaningful benefit in advanced melanoma, including therapeutically challenging PD-1–refractory settings.

Methodologically, the analysis employed a Bayesian random-effects framework on the logit scale to stabilize variance in proportional outcomes and account for anticipated between-study heterogeneity. The estimated heterogeneity parameter (τ ≈ 0.46) reflected substantial variability in trial design, patient selection, treatment combinations, and outcome assessment across included studies. Compared with conventional I² estimates, which may become unstable in sparse or clinically heterogeneous datasets, τ provides a more interpretable measure of dispersion on the modeled scale [24].

An additional advantage of the Bayesian framework is its ability to generate full posterior distributions, thereby supporting probabilistic interpretation and transparent uncertainty quantification in the setting of emerging and heterogeneous immunotherapy evidence. Nevertheless, the analysis assumes exchangeability across studies and cannot eliminate indirectness arising from the absence of head-to-head comparative trials. Consequently, conclusions regarding relative therapeutic superiority between TIL and engineered viral vector strategies remain limited and should be interpreted cautiously.

Importantly, the findings may support complementary rather than competing roles for these immunotherapeutic approaches within evolving melanoma treatment paradigms. Viral vector therapies may enhance antigen presentation and immune priming within immunologically “cold” tumors, whereas

TIL therapy provides highly personalized adoptive cellular responses capable of overcoming established immune resistance. This raises the possibility that biomarker-guided sequencing or combinatorial integration of viral priming followed by adoptive cellular therapy could represent a promising future strategy for optimizing durable antitumor immunity in advanced melanoma.

### Strengths and Limitations

A major strength of this review is the structured synthesis of both randomized and prospective single-arm studies, allowing broader clinical representation across heterogeneous melanoma treatment settings. Included studies captured diverse therapeutic contexts, including PD-1–exposed and checkpoint-refractory populations, Seitter et al. [18], while also reflecting important feasibility and toxicity considerations associated with TIL delivery, such as interleukin-2–supported regimens [17]. Stratification by therapeutic modality, PD-1 exposure, and combination treatment status further improved the interpretability of pooled findings. In addition, structured risk-of-bias assessment using RoB 2 and ROBINS-I together with outcome-level certainty grading through GRADE enhanced methodological transparency and interpretive rigor.

From a quantitative perspective, logit transformation reduced instability associated with proportional outcome data, while Bayesian random-effects modeling accounted for anticipated between-study heterogeneity. The primary Bayesian synthesis included eight studies with sufficiently comparable ORR reporting, while all eligible studies were retained in the qualitative synthesis. Exploratory sensitivity analyses and meta-regression were additionally performed to evaluate the robustness of pooled estimates and assess potential effect modifiers, although these analyses were interpreted as supportive rather than causal inferences.

Several limitations should also be acknowledged. First, no randomized head-to-head trials directly comparing TIL and engineered viral vector therapies were identified, rendering comparisons indirect and susceptible to confounding arising from differences in baseline risk, prior treatment exposure, eligibility criteria, and therapeutic context, including monotherapy versus checkpoint inhibitor combinations. Second, although ORR was consistently reported and therefore suitable for quantitative pooling, progression-free survival, overall survival, and toxicity outcomes were variably defined and inconsistently reported across studies, limiting synthesis of long-term therapeutic durability and safety.

Third, study screening and data extraction were conducted by a single reviewer, introducing potential selection and extraction bias despite the use of predefined eligibility criteria and standardized extraction procedures. Finally, generalizability may be constrained by the geographic concentration and demographic composition of included cohorts, including the presence of a Japan-based all-male study population. Small-study effects were evaluated using standard-error-based funnel plots and Egger’s testing; however, the ability to detect publication bias remains limited in the setting of modest study numbers and substantial heterogeneity.

### Comparison with Previous Work and Limitations of Prior Studies

Previous systematic reviews of tumor-infiltrating lymphocyte (TIL) therapy in melanoma have reported pooled objective response rates (ORRs) broadly ranging between 34% and 44% [23]. Similarly, clinical studies evaluating oncolytic virotherapy, including herpes simplex virus (HSV)-based platforms such as talimogene laherparepvec (T-VEC), have demonstrated variable response rates, with more favorable responses frequently observed in combination with immune checkpoint inhibitors [12–14, 19, 21].

More recent syntheses evaluating immune checkpoint blockade, including CTLA-4, PD-1, and LAG-3–targeted regimens, continue to demonstrate improved survival outcomes with combination immunotherapy approaches while also highlighting persistent primary and acquired resistance in a substantial proportion of patients [25]. This ongoing therapeutic limitation provides a strong clinical rationale for the continued development of alternative and complementary immunotherapeutic strategies, including adoptive cellular therapies and engineered viral vector platforms.

Importantly, earlier evidence syntheses have largely evaluated TIL and viral vector therapies independently, with limited comparative contextualization across mechanistically distinct immunotherapeutic modalities. Many prior analyses also relied heavily on single-arm studies and conventional frequentist pooling approaches, which may produce unstable interval estimates when synthesizing proportional outcomes from sparse or clinically heterogeneous datasets [26, 27]. In addition, relatively few prior reviews stratified findings according to clinically important modifiers such as PD-1 exposure status, treatment sequencing, or checkpoint inhibitor combination context.

By integrating both TIL and engineered viral vector immunotherapies within a unified systematic review and supplementing the synthesis with Bayesian random-effects modeling and exploratory meta-regression, the present study provides a probabilistic comparative framework for evaluating response outcomes across heterogeneous melanoma trial designs. Collectively, these findings support the continued parallel development of both therapeutic platforms and reinforce the potential importance of biomarker-guided sequencing and combinatorial immunotherapy strategies in advanced melanoma.

### Implications and Future Directions

This systematic review supplemented by Bayesian meta-analysis addressed a major evidence gap in melanoma immunotherapy research: the absence of direct comparative synthesis between tumor-infiltrating lymphocyte (TIL) therapies and engineered viral vector immunotherapies. In the absence of head-to-head randomized trials, the present analysis necessarily integrated heterogeneous prospective and single-arm studies using a Bayesian random-effects framework to estimate pooled therapeutic response and characterize uncertainty across diverse treatment contexts. Although this probabilistic approach supports synthesis in sparse and clinically heterogeneous evidence settings, it cannot fully account for confounding arising from differences in trial design, patient selection, treatment combinations, or endpoint assessment.

Accordingly, future research should prioritize prospective comparative trial designs capable of directly evaluating the relative and potentially complementary roles of these immunotherapeutic strategies. Based on the heterogeneity identified in the present analysis together with emerging mechanistic rationale from translational and preclinical studies, an exploratory three-arm Phase IIb randomized framework is proposed (Appendix A1). This design would compare: (1) TIL monotherapy, (2) engineered viral vector monotherapy, and (3) sequential viral priming followed by TIL infusion. Such an approach would permit evaluation of both relative efficacy and potential therapeutic synergy while controlling for adjunctive checkpoint inhibition and enabling harmonized outcome assessment across treatment groups.

Biomarker-guided stratification should also represent a central component of future melanoma immunotherapy trials. Although several included studies referenced biomarkers such as PD-L1 expression, tumor mutational burden (TMB), and intratumoral T-cell density, few prospectively incorporated these variables into treatment allocation or stratified analysis. Integration of predictive biomarkers, including IFN-γ–associated gene signatures, baseline CD8⁺ T-cell infiltration, and RNA-based immune classifiers, may improve patient selection and therapeutic sequencing, particularly within clinically challenging PD-1–refractory populations.

Standardization of clinical outcomes represents an additional priority for future evidence generation. The present synthesis was constrained by inconsistent reporting and heterogeneous definitions of ORR, progression-free survival (PFS), overall survival (OS), and treatment-related toxicity across studies. Future trials should adopt harmonized core outcome frameworks incorporating conventional oncologic endpoints together with immune-specific response criteria such as iRECIST, in addition to patient-reported quality-of-life and health economic measures. Greater standardization would improve cross-study interpretability and facilitate future individual patient data meta-analyses with enhanced precision and reduced methodological bias.

Important practical barriers to implementation must also be addressed. TIL therapy remains resource-intensive, requiring specialized manufacturing infrastructure and lymphodepleting preparative regimens, whereas intratumoral viral vector delivery remains anatomically constrained and may be less feasible in disseminated visceral disease. Advances in decentralized cell manufacturing, cryopreservation workflows, scalable expansion platforms, and systemic or targeted viral delivery technologies may therefore be essential for broader clinical accessibility and real-world implementation.

In conclusion, this study provides one of the first probabilistic comparative syntheses of TIL and engineered viral vector immunotherapies in advanced melanoma. While interpretation remains constrained by underlying study heterogeneity and the absence of direct randomized comparisons, the findings support continued parallel development of both therapeutic platforms and reinforce the potential importance of biomarker-guided sequencing approaches. Ultimately, rigorously designed comparative and biomarker-informed trials will be required to determine the optimal integration, sequencing, and personalization of these emerging immunotherapeutic strategies in advanced melanoma.

## Conclusion

This systematic review supplemented by Bayesian meta-analysis evaluated the comparative efficacy, safety, and translational integration potential of tumor-infiltrating lymphocyte (TIL) therapies and engineered viral vector immunotherapies in advanced melanoma. While pooled Bayesian estimates suggested greater standalone response activity for TIL-based approaches, particularly in PD-1–refractory populations, engineered viral vector therapies demonstrated enhanced activity in combination with immune checkpoint inhibition. Collectively, these findings do not establish the superiority of one modality over the other; rather, they support complementary and potentially synergistic roles that may vary according to clinical and immunologic context.

Mechanistically, TIL therapy provides a highly personalized adoptive cellular approach through reinfusion of tumor-reactive lymphocytes, whereas engineered viral vectors act in situ to enhance antigen presentation and stimulate both innate and adaptive antitumor immune responses. This biological distinction supports further investigation of sequencing and combination strategies, including viral immune priming followed by adoptive cellular therapy, particularly in immunologically “cold” or therapeutically refractory melanoma.

Rather than supporting a uniform treatment paradigm, the findings of this review reinforce the importance of individualized immunotherapy selection based on tumor accessibility, prior checkpoint inhibitor exposure, immune biomarker profiles, and patient fitness. The Bayesian meta-analytic framework additionally enabled probabilistic estimation and transparent uncertainty quantification while accounting for substantial heterogeneity across included trial designs.

Future research should prioritize biomarker-guided randomized studies incorporating harmonized endpoints and prospective comparative frameworks capable of refining therapeutic sequencing and treatment selection. Overall, this review provides both a quantitative synthesis and a translational rationale to support the continued evolution of precision immunotherapy strategies in advanced melanoma.

## Statements and Declarations

### Author contributions

Joshua Anyachor conceived the study, designed the methodology, conducted the literature search, screened and selected eligible studies, extracted the data, performed the statistical analyses, interpreted the findings, and drafted and revised the manuscript. The author approved the final version of the manuscript and accepts responsibility for the integrity and accuracy of the work.

### Use of Artificial Intelligence

AI-assisted tools were used for language refinement and editorial support. All study design decisions, data extraction, statistical analyses, interpretation of findings, and final conclusions were reviewed, verified, and approved by the author, who takes full responsibility for the integrity and accuracy of the manuscript.

## Ethical approval

This article does not contain any studies involving human participants or animals performed directly by the author. All analyses were conducted using previously published data, and no additional ethical approval was required. The author takes full responsibility for the study design, data interpretation, and conclusions presented in this work.

## Consent to Participate

Not applicable

## Consent to Publish

Not applicable

## Data Availability Statement

All data analyzed in this study were derived from previously published articles included in the systematic review. Analytical methods, extracted summary data, and supplementary materials are provided within the manuscript and appendices.

## Data Availability

All data analyzed in this study were derived from previously published articles, clinical trial reports, and supplementary materials included in the systematic review. Extracted summary data, analytical methods, and code used for the Bayesian meta-analysis and exploratory meta-regression are provided within the manuscript and appendices. No individual participant-level data or restricted-access datasets were used.

## Appendix

**Appendix A1.** Proposed Trial: TIL vs. Viral vs. Combination in Advanced Melanoma.

| Element | Description |
| --- | --- |
| Study Design | Phase 2b, multi-center, randomized controlled trial |
| Population | Adults $\geq 18$ with unresectable stage III/IV melanoma, refractory to anti-PD-1 |
| Arms (3 total) | 1. TIL therapy alone<br>2. Viral vector therapy alone<br>3. Viral priming followed by TIL |
| Randomization | 1:1:1, stratified by PD-L1 status and tumor burden |
| Primary Endpoint | Objective Response Rate (RECIST v1.1) |
| Secondary Endpoints | PFS, OS, duration of response, safety (grade $\geq 3$ AEs), quality of life |
| Inclusion Criteria | Aged $\geq 18$, ECOG 0–1, prior ICI failure, injectable or resectable lesions |
| Biomarker Subgroups | PD-L1, TMB, IFN-γ signature (for stratified and exploratory analysis) |
| Statistical Plan | Frequentist parallel-group design; intention-to-treat analysis |

*Note: This Phase 2b trial includes three arms TIL, viral vector, and combination therapy. Eligible patients are aged $\geq 18$ with advanced melanoma, including those post-PD-1 treatment. Biomarker stratification and standardized endpoints are included to enhance generalizability. Sample size will depend on funding and feasibility.*

## METHODOLOGICAL CONTRIBUTION TO SYNTHESIS

Table A2 summarizes key methodological limitations and their influence on study interpretation and synthesis.

**Table A2.**
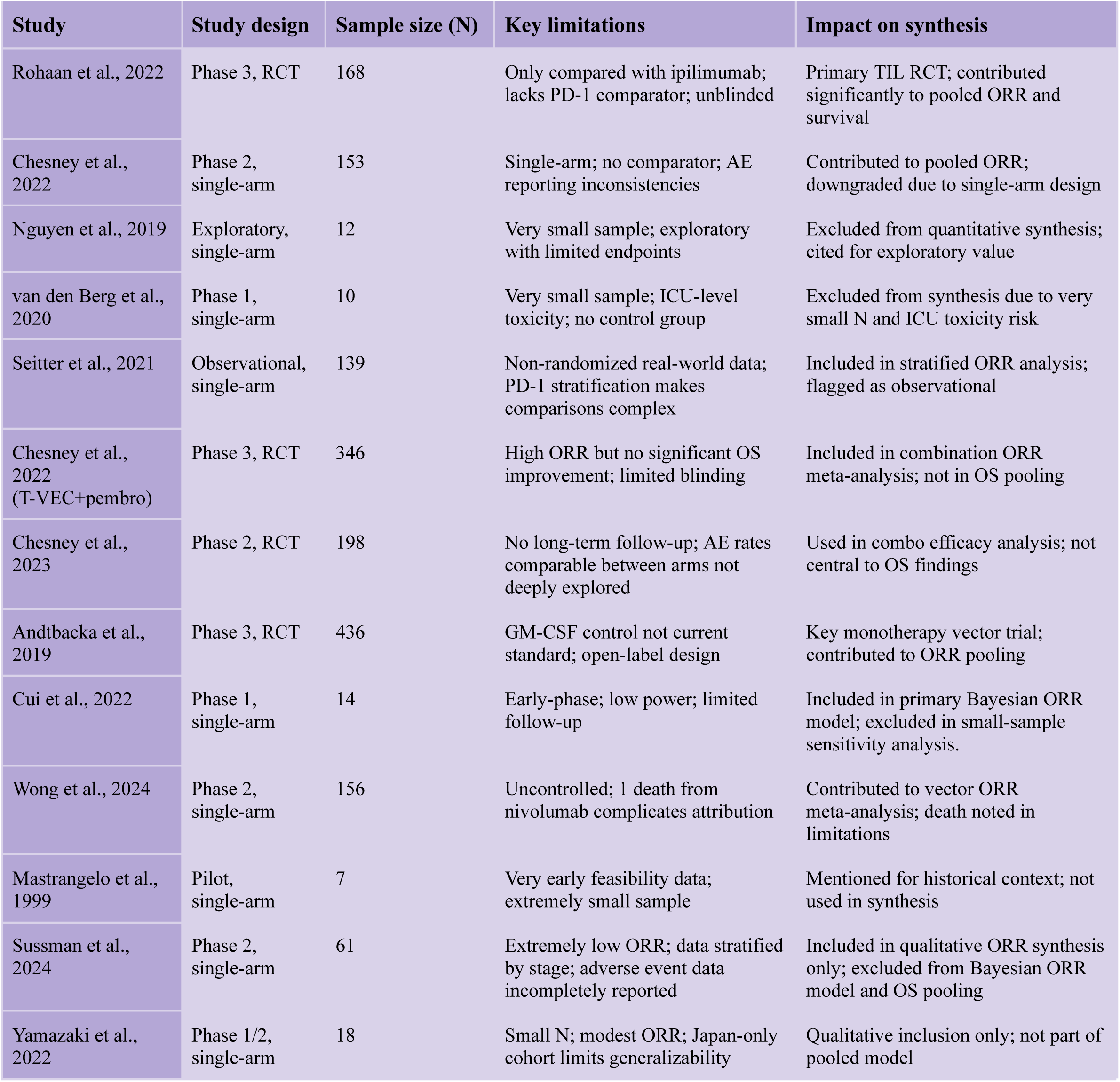
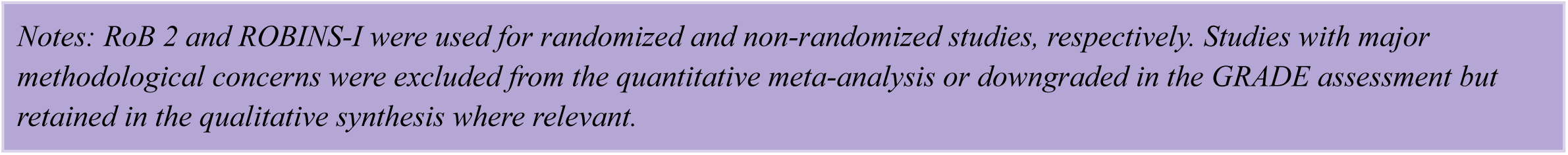
Key Limitations and Contribution to Synthesis. Summary of study level methodological issues and their role in inclusion, exclusion, or weighting in the meta-analysis

| Study | Study design | Sample size (N) | Key limitations | Impact on synthesis |
| --- | --- | --- | --- | --- |
| Rohaani et al., 2022 | Phase 3, RCT | 168 | Only compared with ipilimumab; lacks PD-1 comparator; unblinded | Primary TIL RCT; contributed significantly to pooled ORR and survival |
| Chesney et al., 2022 | Phase 2, single-arm | 153 | Single-arm; no comparator; AE reporting inconsistencies | Contributed to pooled ORR; downgraded due to single-arm design |
| Nguyen et al., 2019 | Exploratory, single-arm | 12 | Very small sample; exploratory with limited endpoints | Excluded from quantitative synthesis; cited for exploratory value |
| van den Berg et al., 2020 | Phase 1, single-arm | 10 | Very small sample; ICU-level toxicity; no control group | Excluded from synthesis due to very small N and ICU toxicity risk |
| Seitter et al., 2021 | Observational, single-arm | 139 | Non-randomized real-world data; PD-1 stratification makes comparisons complex | Included in stratified ORR analysis; flagged as observational |
| Chesney et al., 2022 (T-VEC+pembro) | Phase 3, RCT | 346 | High ORR but no significant OS improvement; limited blinding | Included in combination ORR meta-analysis; not in OS pooling |
| Chesney et al., 2023 | Phase 2, RCT | 198 | No long-term follow-up; AE rates comparable between arms not deeply explored | Used in combo efficacy analysis; not central to OS findings |
| Andtbacka et al., 2019 | Phase 3, RCT | 436 | GM-CSF control not current standard; open-label design | Key monotherapy vector trial; contributed to ORR pooling |
| Cui et al., 2022 | Phase 1, single-arm | 14 | Early-phase; low power; limited follow-up | Included in primary Bayesian ORR model; excluded in small-sample sensitivity analysis. |
| Wong et al., 2024 | Phase 2, single-arm | 156 | Uncontrolled; 1 death from nivolumab complicates attribution | Contributed to vector ORR meta-analysis; death noted in limitations |
| Mastrangelo et al., 1999 | Pilot, single-arm | 7 | Very early feasibility data; extremely small sample | Mentioned for historical context; not used in synthesis |
| Sussman et al., 2024 | Phase 2, single-arm | 61 | Extremely low ORR; data stratified by stage; adverse event data incompletely reported | Included in qualitative ORR synthesis only; excluded from Bayesian ORR model and OS pooling |
| Yamazaki et al., 2022 | Phase 1/2, single-arm | 18 | Small N; modest ORR; Japan-only cohort limits generalizability | Qualitative inclusion only; not part of pooled model |

*Notes: RoB 2 and ROBINS-I were used for randomized and non-randomized studies, respectively. Studies with major methodological concerns were excluded from the quantitative meta-analysis or downgraded in the GRADE assessment but retained in the qualitative synthesis where relevant.*

**Appendix Table A3.** Quality Assessment of RCTs Using RoB 2.

| Study | Randomization Process | Deviations from Intended Interventions | Missing Outcome Data | Measurement of Outcome | Selection of Reported Result | Overall Risk |
| --- | --- | --- | --- | --- | --- | --- |
| Rohaani et al. (2022) | Low – Randomized, controlled phase III trial. | Low – Blinding minimizes deviations. | Low – Minimal missing data, well addressed. | Low – Blinded assessment reduces bias. | Low – Pre-specified endpoints reported fully. | Low – Rigorous methodology with blinding. |
| Chesney et al. (2022) | Low – Double-blind, placebo-controlled trial. | Low – Blinding minimizes deviations. | some – Minimal attrition, well documented. | Low – Blinded assessment reduces bias. | some – Pre-specified and transparently reported. | some – Rigorous methodology with blinding. |
| Chesney et al. (2023) | Low – Randomized design reported. | Low – Blinding minimizes deviations. | Low – Well tracked and reported. | Low – Blinded assessment reduces bias. | Low – Consistent, clearly defined outcomes. | Low – Rigorous methodology with blinding. |
| Andtbacka et al. (2019) | Low – Adequate randomization procedure reported. | Low – Blinding minimizes deviations. | Low – Dropouts well managed. | Low – Blinded assessment reduces bias. | Low – Outcomes reported transparently. | Low – Rigorous methodology with blinding. |

*Note: Each ROB 2 domain includes rating and specific justification directly within the table cells for clarity and space efficiency.*

**Appendix Table A4.**
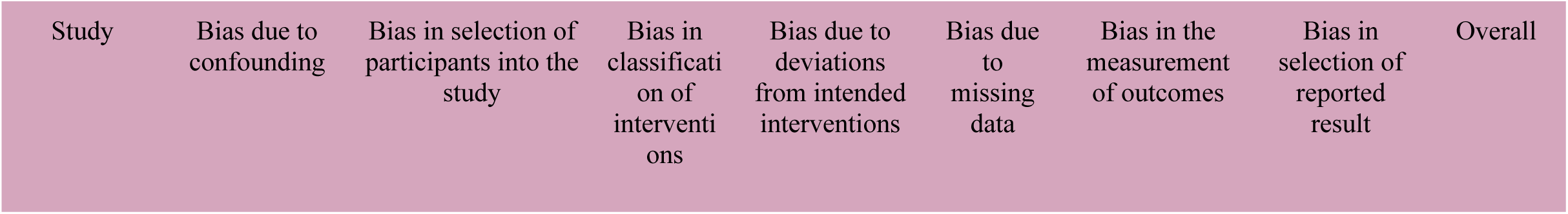
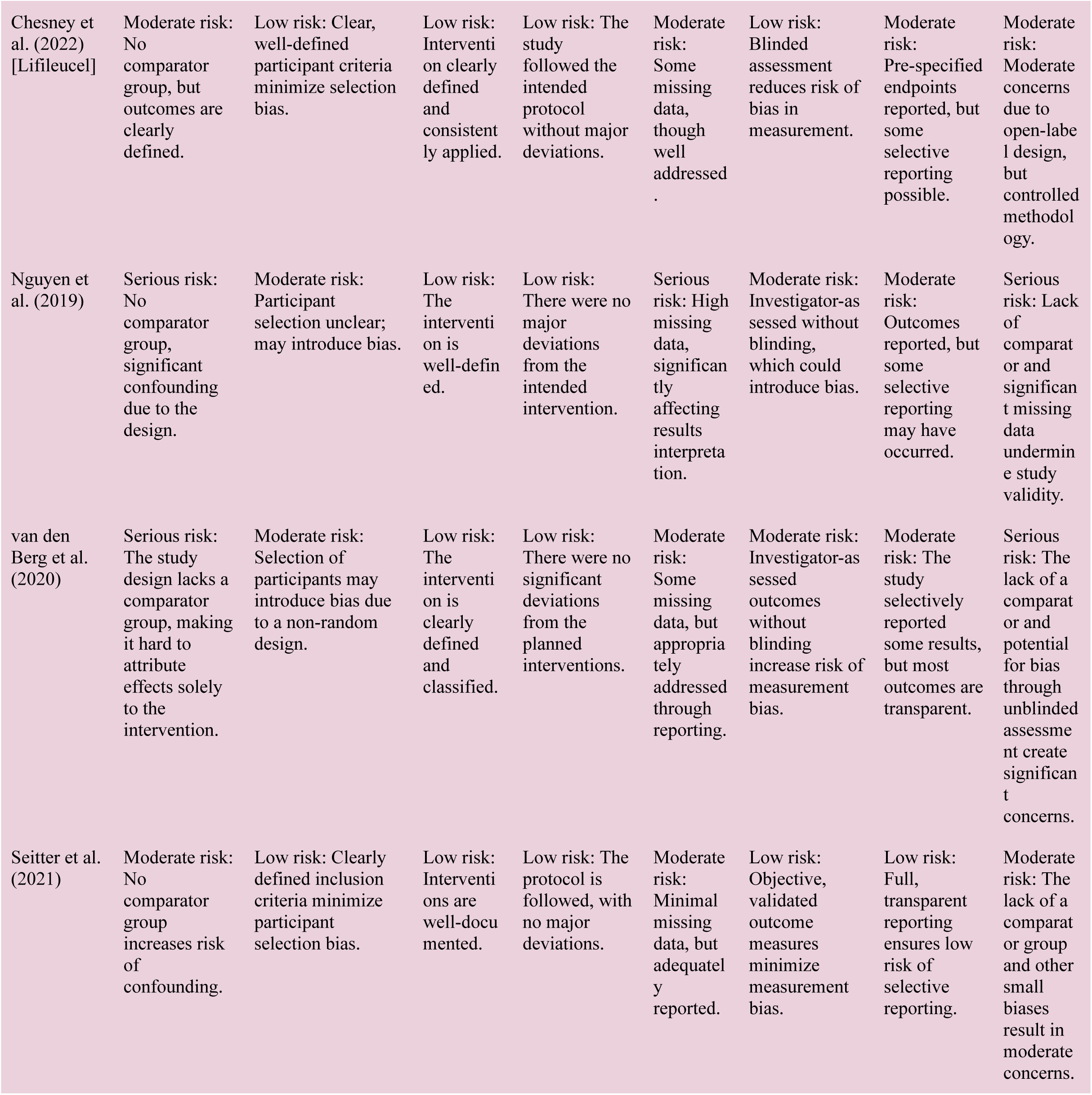
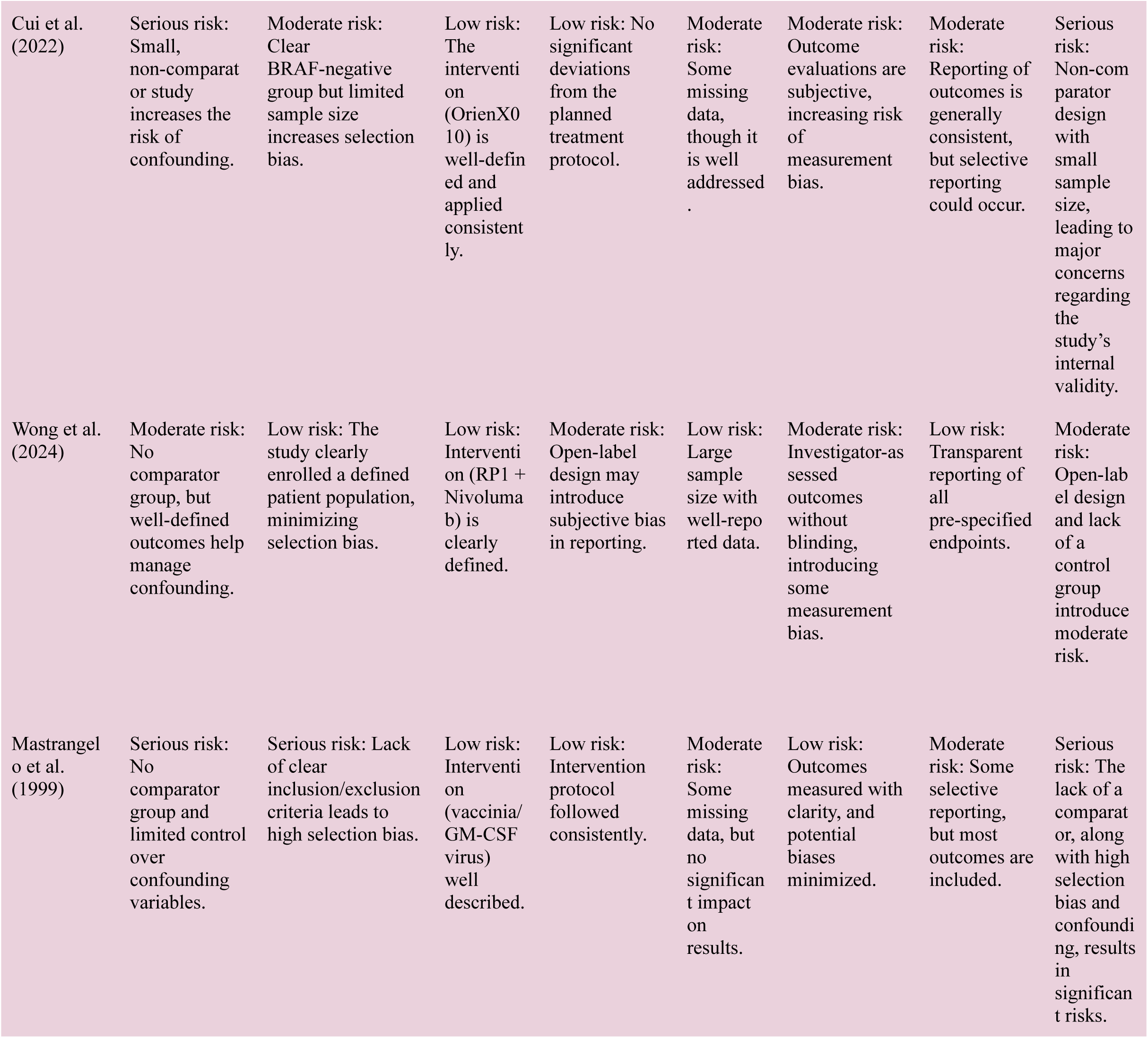
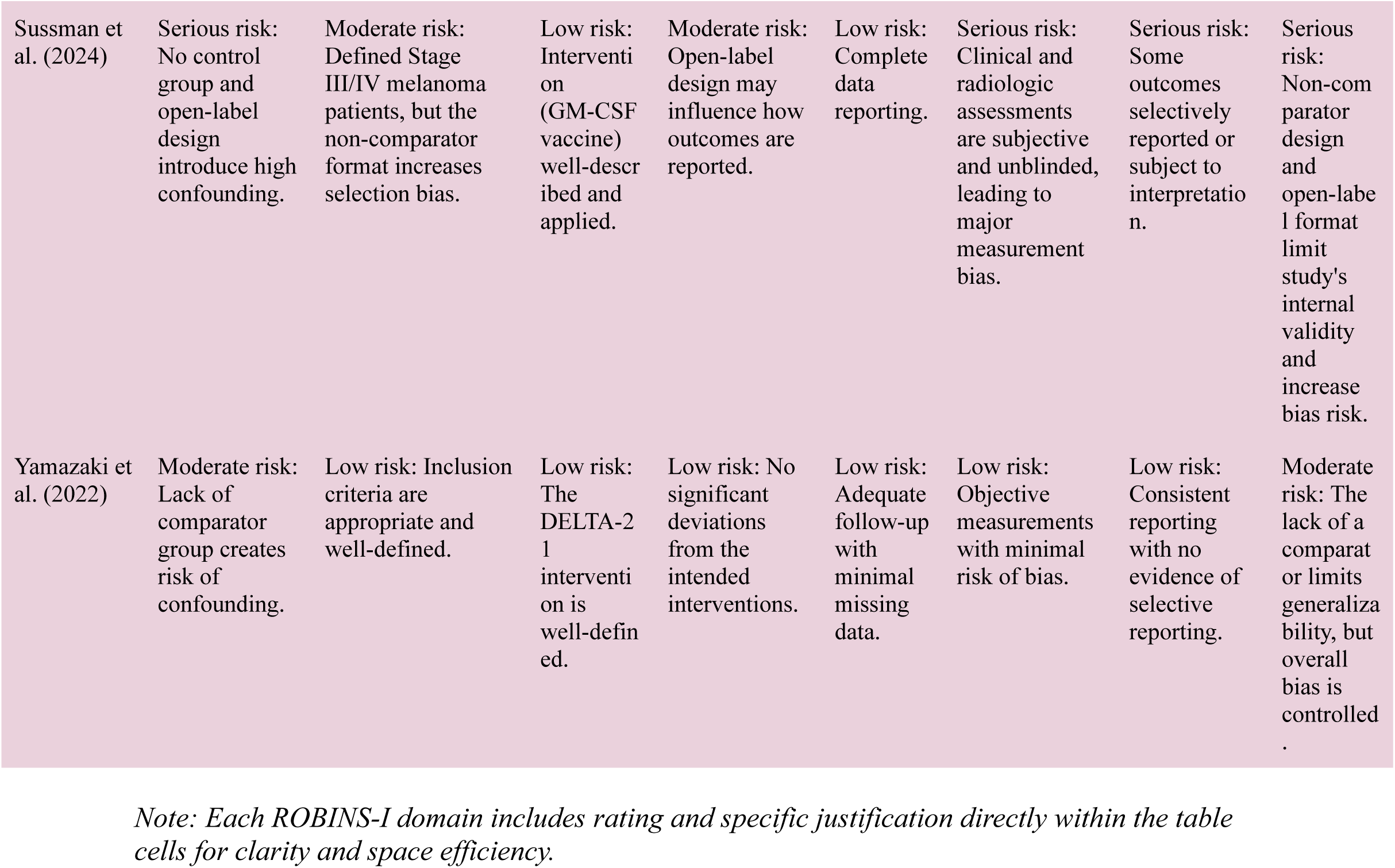
Quality Assessment of Single-Arm Studies using ROBINS-1.

**Appendix A5.**
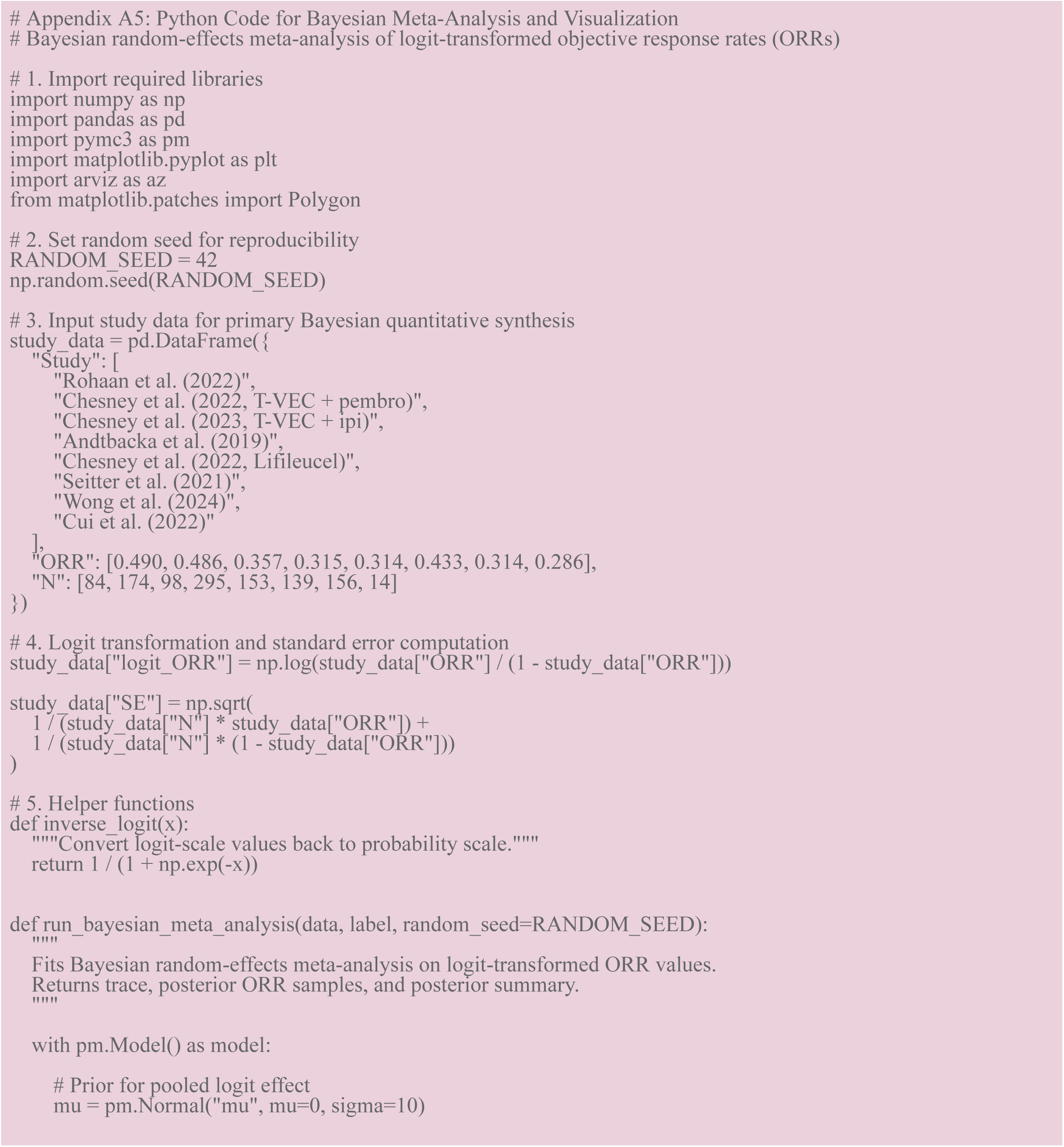
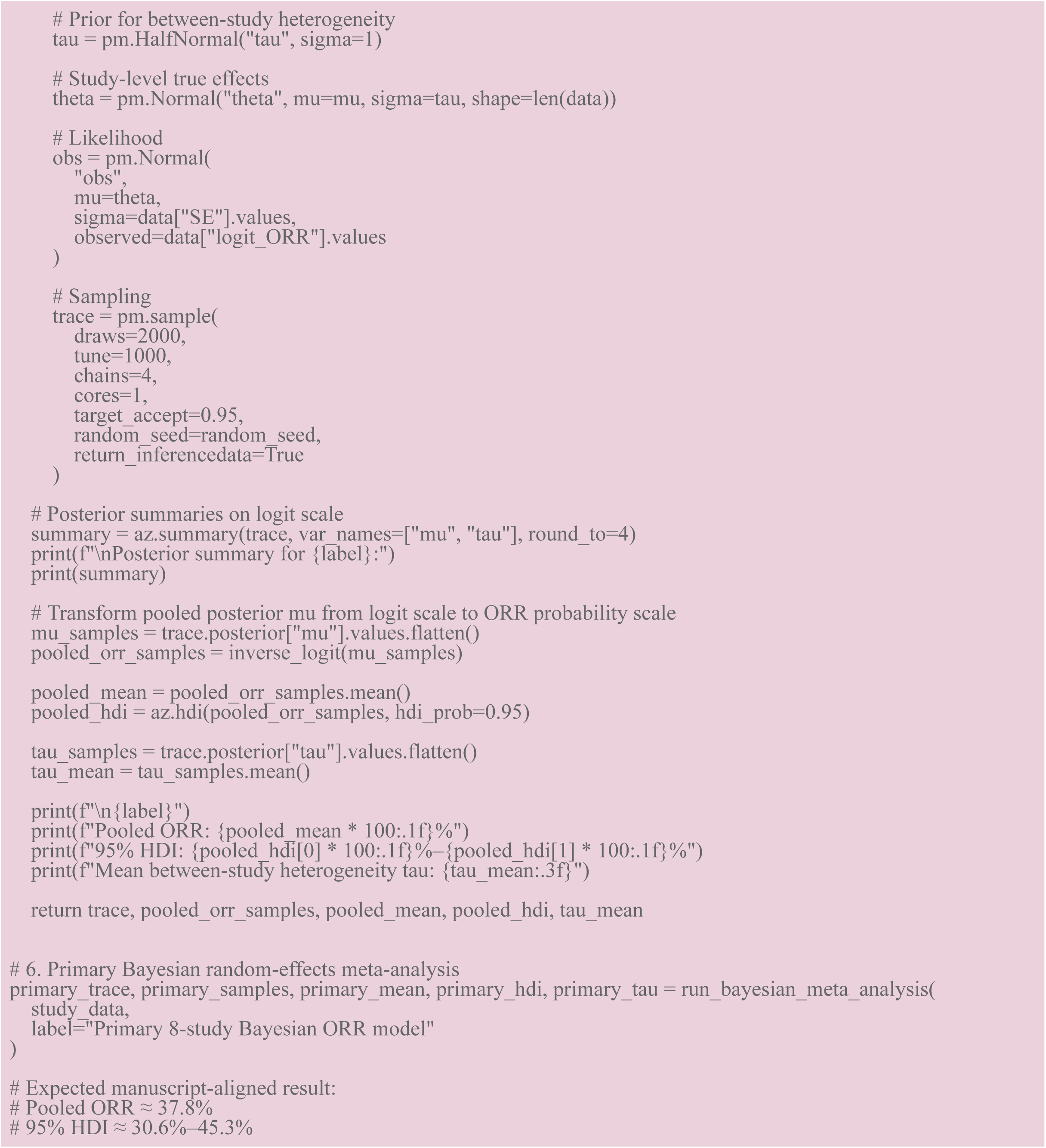
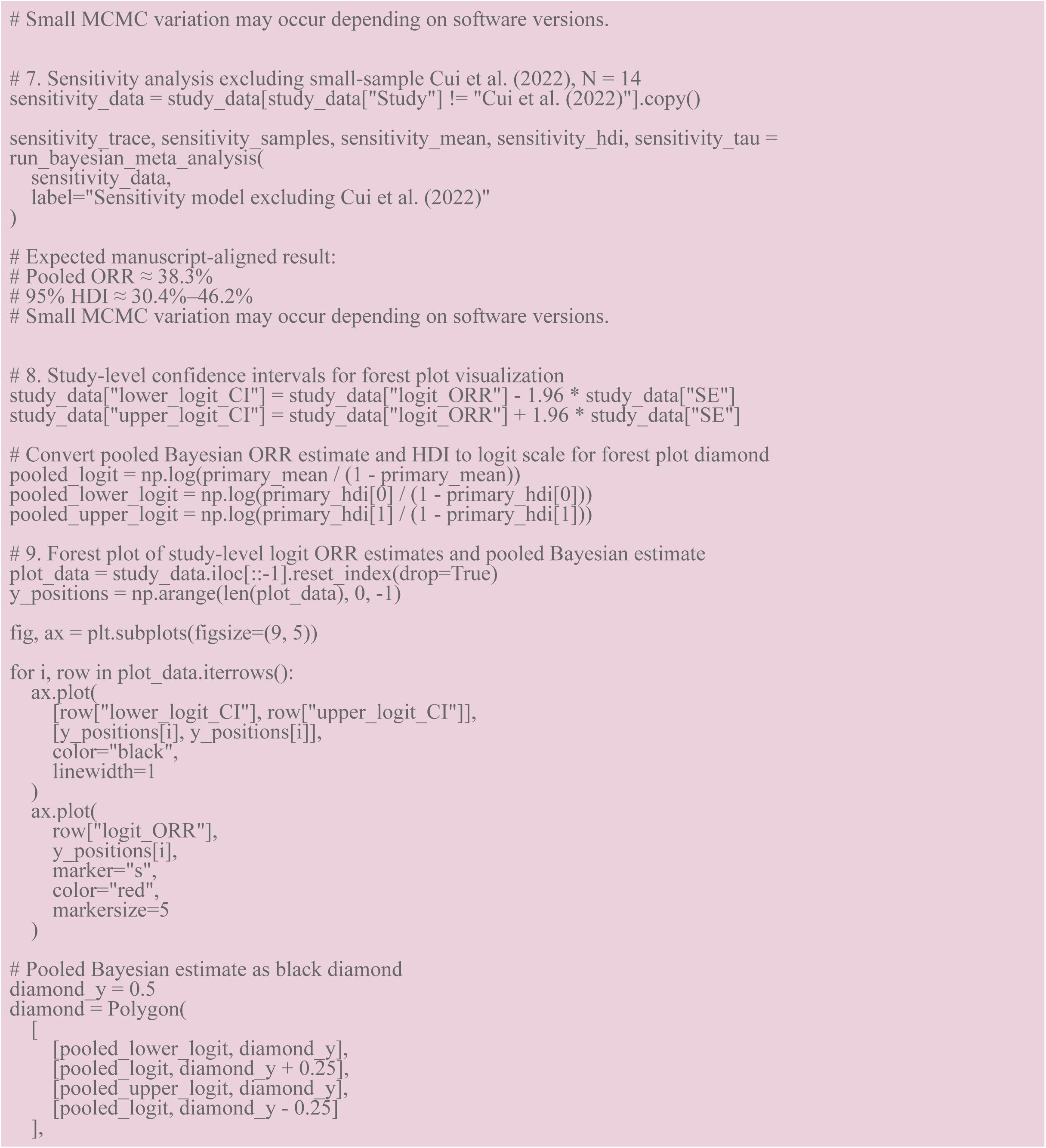
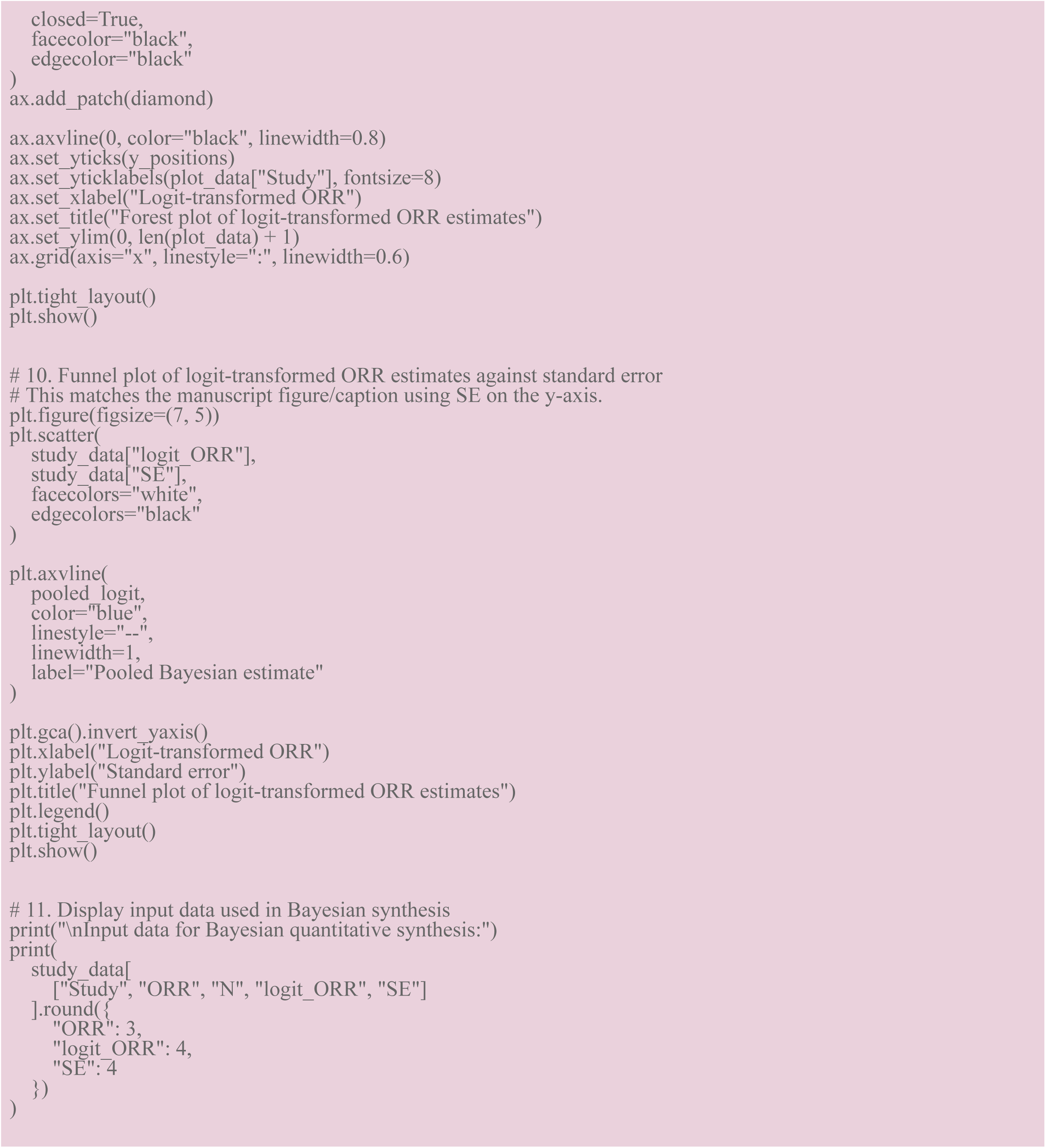
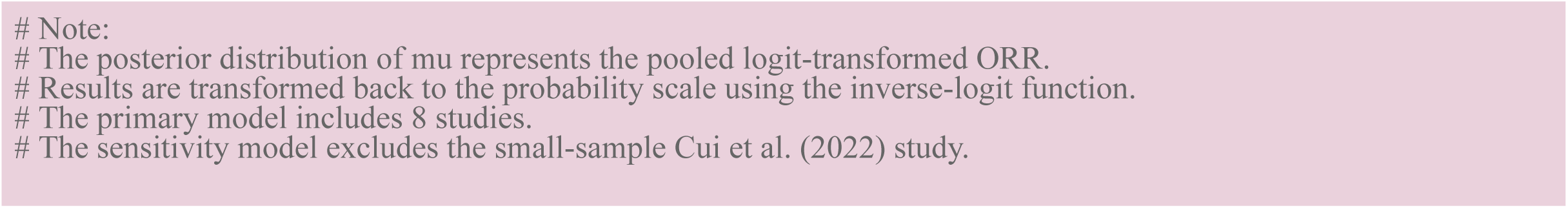
Python Code for Bayesian Meta-Analysis and Visualization.

**Appendix A6.**
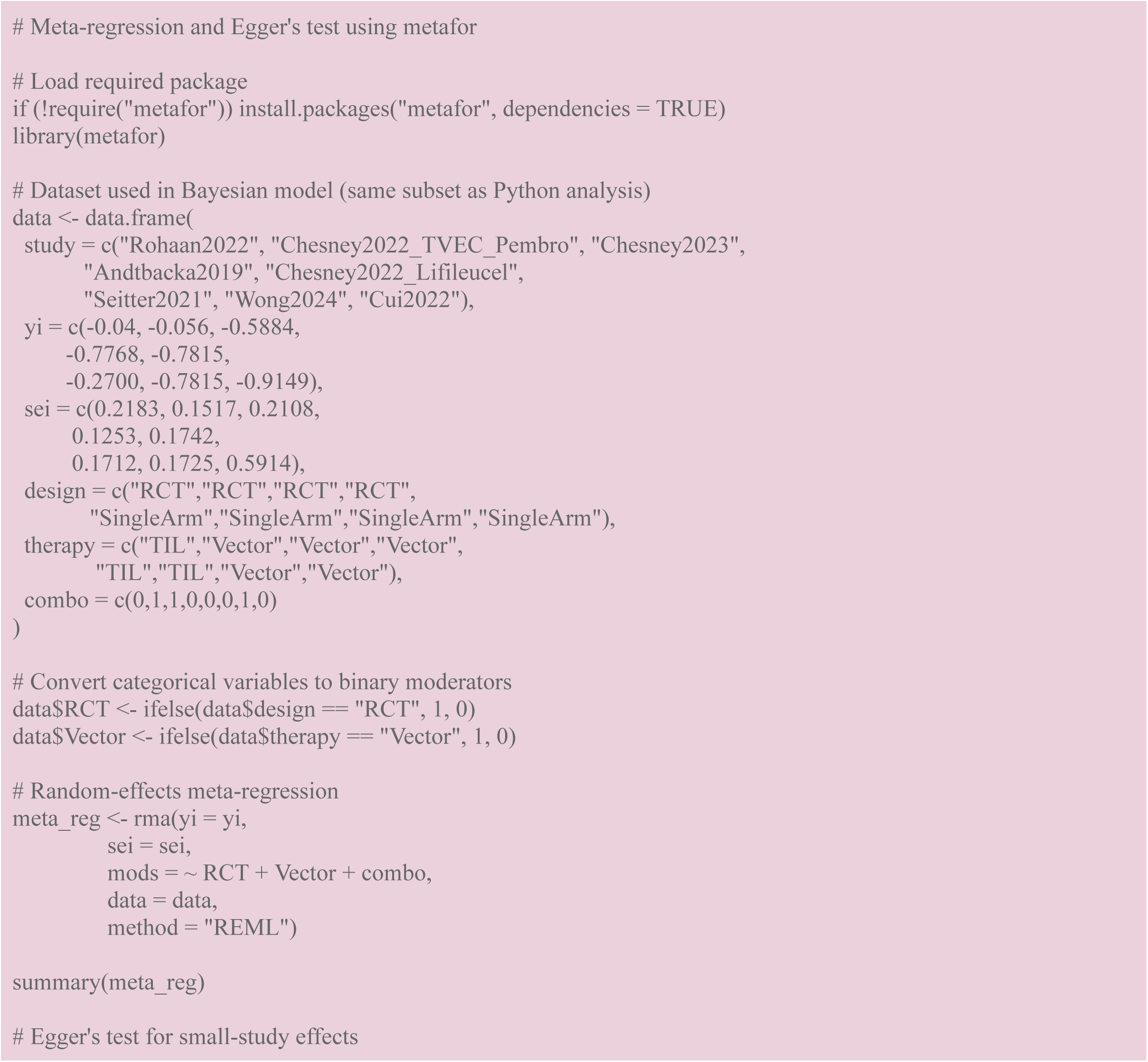
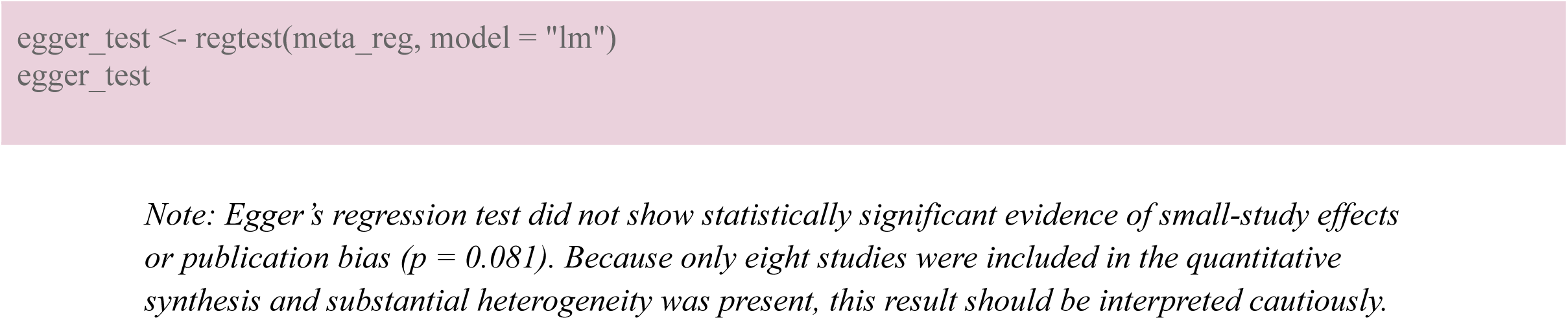
Meta-Regression and Egger’s Test R Code Using metafor.

**Appendix A7.**
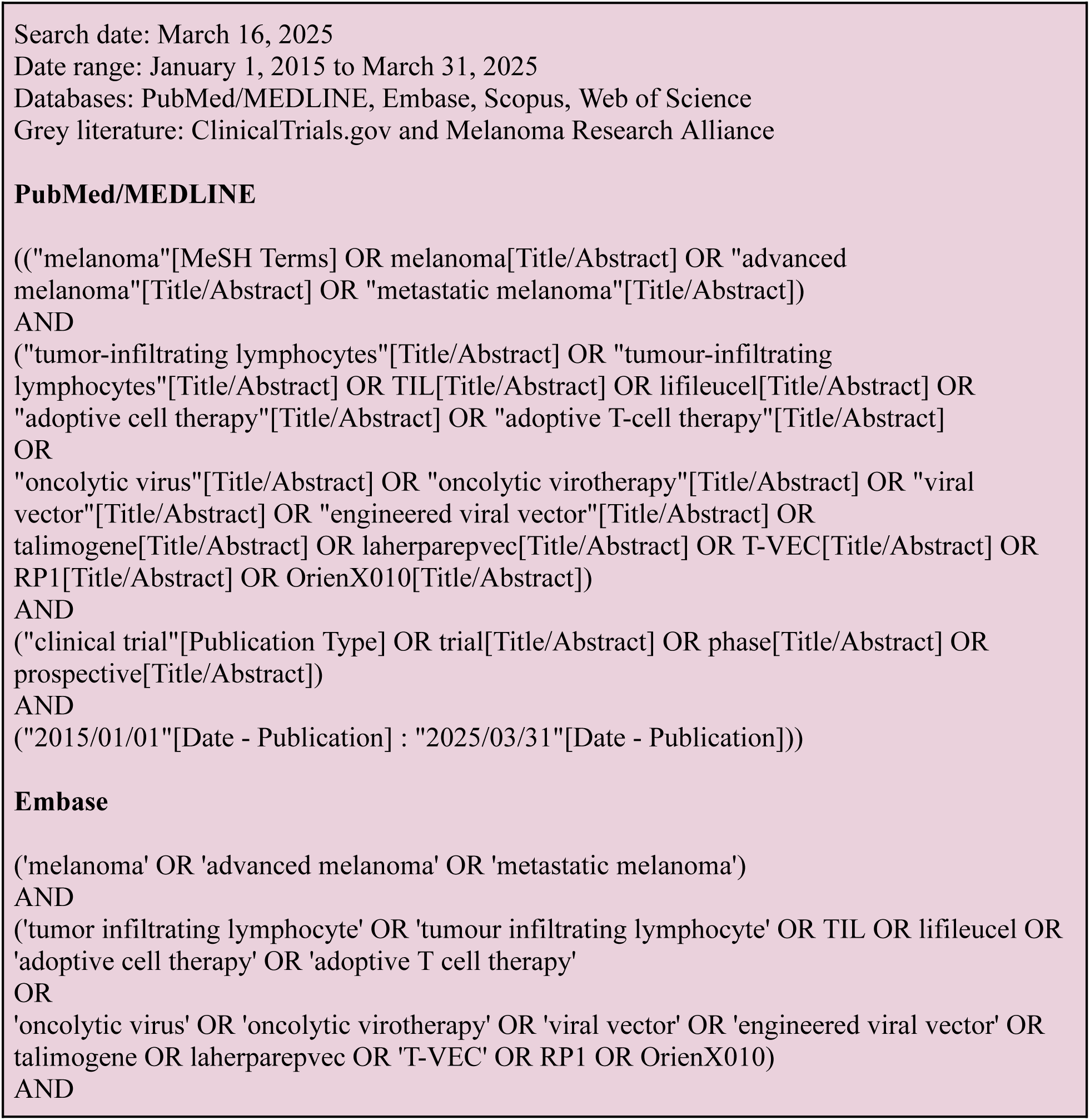
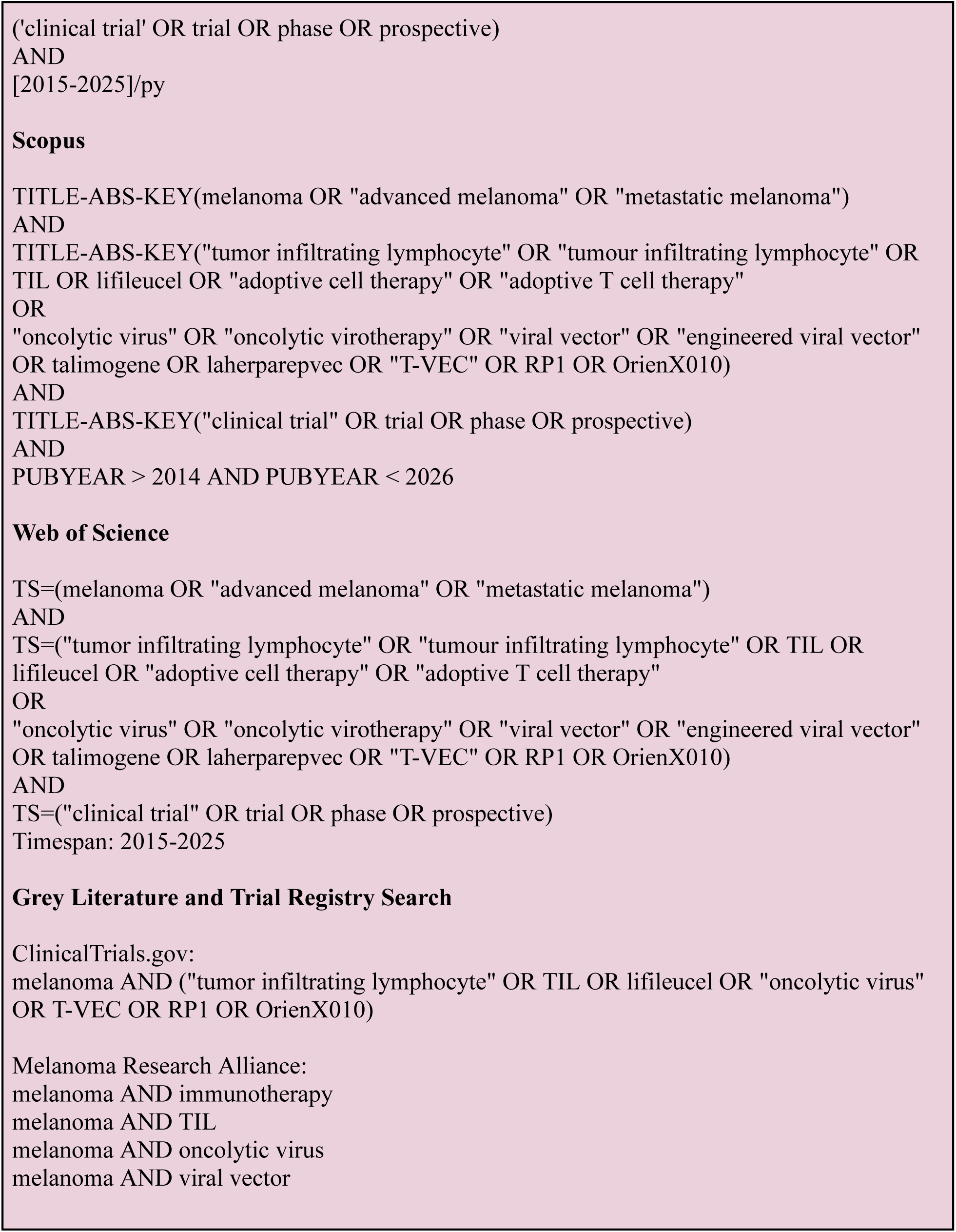
Full Database Search Strategy.

